# A proteomic signature of healthspan

**DOI:** 10.1101/2024.06.26.24309530

**Authors:** Chia-Ling Kuo, Peiran Liu, Gabin Drouard, Eero Vuoksimaa, Jaakko Kaprio, Miina Ollikainen, Zhiduo Chen, Luke C. Pilling, Janice L. Atkins, Richard H. Fortinsky, George A. Kuchel, Breno S. Diniz

## Abstract

The focus of aging research has shifted from increasing lifespan to enhancing healthspan to reduce the time spent living with disability. Despite significant efforts to develop biomarkers of aging, few studies have focused on biomarkers of healthspan. We developed a proteomics-based signature of healthspan (healthspan proteomic score (HPS)) using proteomic data from the Olink Explore 3072 assay in the UK Biobank Pharma Proteomics Project (53,018 individuals and 2920 proteins). A lower HPS was associated with higher mortality risk and several age-related conditions, such as COPD, diabetes, heart failure, cancer, myocardial infarction, dementia, and stroke. HPS showed superior predictive accuracy for these outcomes compared to other biological age measures. Proteins associated with HPS were enriched in hallmark pathways such as immune response, inflammation, cellular signaling, and metabolic regulation. The external validity was evaluated using the Essential Hypertension Epigenetics study with proteomic data also from the Olink Explore 3072 and complementary epigenetic data, making it a valuable tool for assessing healthspan and as a potential surrogate marker to complement existing proteomic and epigenetic biological age measures in geroscience-guided studies.

**Significance:** Despite substantial efforts to develop biomarkers of aging, few studies have focused on biomarkers of healthspan. The challenge lies in the need for long follow-up periods and large sample sizes of healthy individuals to observe aging outcomes. Therefore, developing surrogate biomarkers that can predict healthspan is crucial. We addressed this by developing a proteomics-based signature of healthspan, termed the Healthspan Proteomic Score (HPS), in a healthy cohort. We demonstrated its clinical, predictive, and biological validity in the UK Biobank and Essential Hypertension Epigenetics study, which represents a focused subset of the Finnish Twin Cohort. The HPS, serving as a surrogate marker of healthspan, is useful for gauging an individual’s biological health and monitoring the impact of geroscience-guided interventions.

## Main

Over recent decades, global life expectancy has significantly increased, especially in developing countries. However, the corresponding healthy life expectancy, defined as years lived in full health since birth, has failed to keep up the pace (1). The gap between healthspan and lifespan highlights a substantial challenge posed by chronic diseases among older adults (2), emphasizing the need for novel approaches to narrow the gap and prolong disease-free longevity (3). The geroscience approach is a potential solution, addressing the hypothesis that targeting the hallmarks of aging, rather than individual diseases of aging, may prevent or delay the onset of multiple age-related conditions (4).

Unlike lifespan, which has a universal definition, there is no consensus on the definition of healthspan (5, 6). Previous research has suggested characterizing healthy aging in five domains: physical capability, cognitive function, physiological and musculoskeletal, endocrine, and immune functions (5, 7). For practical purposes, healthspan typically refers to the period of life spent in good health, free from chronic diseases and disabilities of aging (6). Studies aiming to evaluate the effects of interventions on healthspan are challenging due to the need for long follow-up lengths and large sample sizes of healthy individuals to observe the outcomes of interest. Thus, developing surrogate biomarkers that can predict healthspan is crucial for improving the feasibility of clinical trials aimed at testing interventions to prolong healthspan and lifespan.

Composite biomarkers incorporating multiple measures are more robust in predicting age-related outcomes than single biomarkers (8–10). Several composite biomarkers for predicting lifespan or mortality have been developed using clinical biomarkers (11, 12) or omics data (11, 13–15). However, no composite biomarker measures have been developed based on a healthspan definition. To mitigate this gap, we developed a proteomics-based healthspan biomarker (healthspan proteomic score, HPS) using chronological age and expression data of 2,920 proteins at the UK Biobank (UKB) baseline/recruitment (2006–10). In line with previous studies (16, 17), we defined healthspan as the number of years from birth without a major chronic medical condition, including cancer (excluding non-melanoma skin cancer), diabetes (type I diabetes, type II diabetes, and malnutrition-related diabetes), heart failure, myocardial infarction (MI), stroke, chronic obstructive pulmonary disease (COPD), dementia, or death.

Participants from the UK Biobank Pharma Proteomics Project (UKB PPP) were followed up for a mean duration of 13.5 years. Using an independent internal cohort and the Essential Hypertension Epigenetics (EH-Epi) study, HPS was tested against various biological age measures, including proteomic and epigenetic aging clocks. In addition, we investigated the biological processes associated with healthspan through gene set enrichment analysis within the UKB.

## Results

### UKB PPP participants (n=53,018) versus UKB baseline cohort participants not included in the UKB PPP (n=449,251)

Of the 502,268 active UKB participants, 53,018 were included in the UKB Pharma Proteomics Project (PPP) and had proteomic data available, as released by the UKB. Baseline characteristics were comparable between UKB PPP participants and non-participants in the UKB baseline cohort (**Supplementary Table 1**). A previous study (18) comparing the UKB to the UK general population found that UKB participants were more likely to be women, older, and from less socioeconomically deprived areas. They also had healthier lifestyles, lower disease prevalence, and reduced mortality rates. Although the UKB is not representative of the UK general population and cannot support population parameter estimation, its substantial variation in exposures allows for the generalization of associations (19).

### UKB PPP participants free from the conditions in the healthspan definition at baseline (n=43,119)

Of participants included in the UKB PPP, 43,119 (81.3%) had not been diagnosed with any condition included in the healthspan definition at baseline (**Supplementary Table 2**). Their mean age was 56 years (SD=8.2, range 39 to 70), and the majority were of European descent (93.8%) and female (55.8%). 34.3% of participants held a college or university degree, and the mean level of material deprivation, as measured by the Townsend deprivation index, was lower than the UK population average (-1.3 vs. 0). Additionally, 43.4% were previous or current smokers, and the mean BMI was 27.1 (SD=4.6). The prevalence of hypertension and hypercholesterolemia was 23.2% and 11.2%, respectively (**Supplementary Table 3**).

### Baseline characteristics of UKB PPP participants who developed any condition in the healthspan definition during follow-up (n=12,427) versus those who remained free from these conditions (n=30,692)

After a mean follow-up of 13.5 years (from baseline to death or last follow-up, whichever occurred first), 12,427 (28.8%) of 43,119 participants, initially free from the conditions in the healthspan definition at baseline, developed at least one such condition. The incidence rates of these diseases were as follows: cancer at 13.2%, MI at 7.3%, diabetes at 4.9%, COPD at 4.1%, and dementia at 2.4%, with an overall mortality rate of 7.6%. The first diagnosis was at the mean age of 66.7 years (SD=8.1). After excluding participants with multiple first-diagnosed conditions in the healthspan definition (n=1,214, 9.8%), cancer was the most common first-diagnosed condition (n=4,882, 44%), followed by MI (n=2,036, 18.2%), diabetes (n=1,455, 13.0%), and then COPD (n=1,131, 10.1%).

Compared to participants who developed any condition in the healthspan definition during follow-up (n=30,692), participants who remained free from the conditions were younger and more likely to be female or non-white. They had better socioeconomic status, healthier lifestyles, and a lower prevalence of hypertension and hypercholesterolemia at baseline (**Supplementary Table 3**).

### Healthspan Proteomic Score (HPS) development using UKB PPP participants without the conditions in the healthspan definition at baseline (training sample, n=30,184)

Participants who did not have any conditions in the healthspan definition at baseline (n=43,119) were randomly split into training (70%, n=30,184) and test sets (30%, n=12,935). During follow-up, 2,296 (7.6%) participants in the training set and 989 (7.6%) participants in the test set died (**Supplementary Figure 1**). The training and test samples shared similar participant characteristics at baseline (**Supplementary Table 4**).

We developed the HPS using the training set data, including 2,920 proteins and chronological age. In the variable selection, we selected chronological age and 86 proteins based on a Least Absolute Shrinkage and Selection Operator (LASSO) penalized Cox regression model for achieving a more parsimonious model with a close to the optimal deviance for predicting healthspan, following the one-standard-error rule (20) (lambda 0.009277) (**Supplementary Table 5**). Then, chronological age and the selected proteins were used to fit a Gompertz model (**Supplementary Table 5**), where we calculated the 10-year risk of developing a first condition in the healthspan definition (*r*_h_), which was used to derive the healthspan proteomic score (HPS) by 1-*r*_h_. The lower the HPS, the higher the risk. Of note, using the test sample, the Cox regression model, including the 86 proteins, yielded a C-statistic of 0.715, while the model that included the proteins and chronological age produced a C-statistic of 0.718. This similarity in C-statistics suggests that chronological age in the HPS plays a minimal role in predicting the healthspan.

### HPS and the number of conditions in the healthspan definition at baseline among UKB PPP participants (n=53,018)

HPS followed a left-skewed distribution in UKB PPP participants without any condition in the healthspan definition (**Figure 1A**). The distribution shifted to the left and showed a right tail as the number of conditions in the healthspan definition increased. The median HPS in those who had developed one or more conditions in the healthspan definition (0.70 [1 condition], 0.49 [2 conditions], 0.36 [3 conditions], 0.17 [4 conditions], 0.07 [5 conditions]) was significantly lower than that (0.84) in those without any condition included in the healthspan definition at baseline (p<0.001). Among participants with a chronic medical condition at baseline (n=8,136), those with cancer showed the highest median HPS (0.78), and those with MI (0.63), diabetes (0.62), or heart failure (0.62) showed the lowest median HPS (**Figure 1B**).

**Figure 1.**
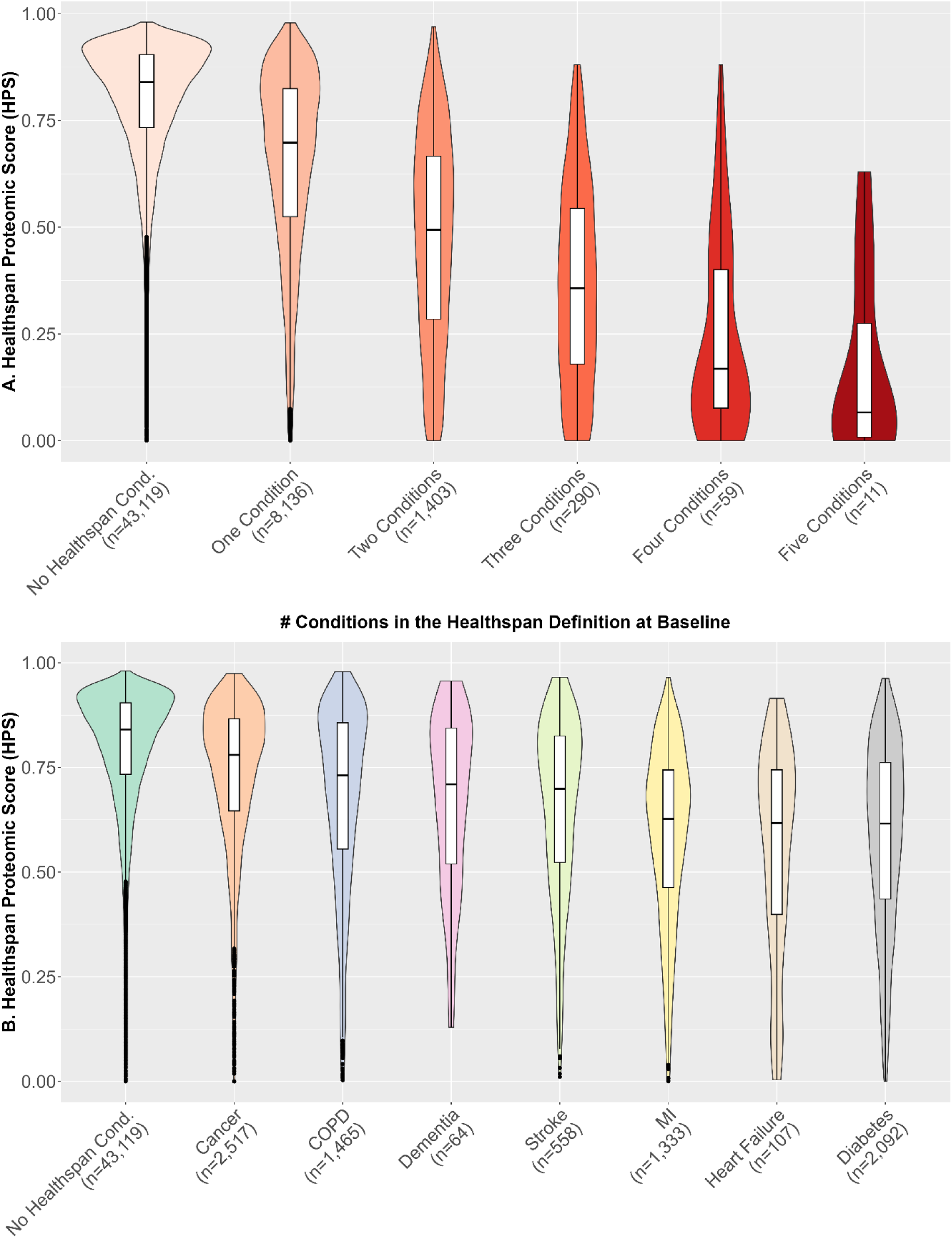
[UKB] Healthspan Proteomic Score (HPS) distributions in UKB PPP participants (n=53,018), separated by the number of conditions in the healthspan definition at baseline (Figure 1A), and within those with a specific condition only at baseline (Figure 1B).

### HPS and disease risk factors at baseline (test sample, n=12,935)

Among the participants in the test set without any condition in the healthspan definition at baseline, the median HPS was significantly lower in males (vs. females), older adults (≥60 years vs. <60 years), former or current smokers (vs. non-smokers), obese participants (BMI≥30 vs. BMI<30), and participants diagnosed with hypertension or hypercholesterolemia before or at baseline (p<0.001, **Supplementary** Figure 2).

### HPS and chronological age, biological age measures, as well as aging traits at baseline (test sample, n=12,935)

HPS showed moderately negative correlations with chronological age (Spearman correlation *r*= -0.73) and with other composite biological age measures (**Supplementary Figure 3**): proteomic aging clock (PAC) (15) (*r*= -0.87), protein-predicted age trained using the elastic net method (ProtAge-EN) (*r*= -0.72) (21), PhenoAge (11) (*r*= -0.79), and BioAge (22) (*r*= -0.74). Moreover, HPS had a weak negative correlation with a 49-item frailty index (23) (*r*= -0.21), based on a wide range of self-reported health deficits, and a weak positive correlation with leukocyte telomere length (24) (*r*= 0.21) (**Supplementary Figure 3**). In addition, HPS was negatively correlated with BMI (*r*= -0.32), systolic blood pressure (*r*= -0.37), and reaction time based on a cognitive function test (*r*= -0.26) (**Supplementary Figure 3**). HPS showed a positive correlation with usual walking pace (*r*= 0.23) and a minimal correlation with maximal grip strength (*r*= -0.01) (**Supplementary Figure 3**).

After regressing out the effect of chronological age, correlations of HPS residuals (HPSRes) with other biological age residuals were significantly reduced (**Supplementary Figure 4**). The correlation between HPSRes and PACRes significantly decreased but remained moderate (*r*= -0.66). HPSRes consistently outperformed PACRes, ProtAge-ENRes, and PhenoAgeRes in correlations with several health variables at baseline, such as BMI, grip strength, and walking pace (**Supplementary Figure 4**).

### HPS at baseline and the risks of developing any or specific conditions in the healthspan definition and other health conditions during follow-up (test sample, n=12,935)

HPS was significantly associated with the development of any condition in the healthspan definition after adjusting for chronological age and other covariates at baseline (false discovery rate [FDR]-adjusted p-value, p_FDR-adj_=8.65×10^-118^). The risk of developing a first condition in the healthspan definition increased as HPS decreased, with an additional 1,600 cases per 0.1 unit decrease in HPS during 100,000 person-years of follow-up (**Table 1**).

**Table 1.** Risk changes associated with a 0.1 unit decrease in HPS for developing a first condition and individual conditions in the healthspan definition, and other medical conditions.

|  | Age-Adjusted |  |  | Fully Adjusted<sup>2</sup> |  |  |  |  |
| --- | --- | --- | --- | --- | --- | --- | --- | --- |
| Outcome | N | Cases | Estimate<sup>1</sup><br>(95% CI) | N | Cases | Estimate<sup>1</sup><br>(95% CI) | P | P<sub>FDR-ADJ</sub> |
| Conditions in the Healthspan Definition |  |  |  |  |  |  |  |  |
| First Cond. in the HS Def. | 12,935 | 3,675 | 1670<br>(1540, 1800) | 12,598 | 3,675 | 1600<br>(1450, 1750) | 4.55E-119 | 8.65E-118 |
| Mortality | 12,935 | 989 | 523<br>(460, 586) | 12,598 | 989 | 524<br>(450, 598) | 7.65E-53 | 7.26E-52 |
| Diabetes | 12,935 | 630 | 361<br>(315, 407) | 12,598 | 630 | 317<br>(264, 370) | 1.06E-33 | 6.69E-33 |
| COPD | 12,935 | 515 | 348<br>(298, 398) | 12,598 | 515 | 344<br>(286, 402) | 1.28E-31 | 4.85E-31 |
| Cancer | 12,935 | 1,685 | 430<br>(365, 495) | 12,598 | 1,685 | 448<br>(370, 526) | 4.12E-31 | 1.30E-30 |
| Heart Failure | 12,935 | 375 | 230<br>(189, 271) | 12,598 | 375 | 230<br>(183, 277) | 5.52E-24 | 1.50E-23 |
| Myocardial Infarction | 12,935 | 931 | 369<br>(314, 424) | 12,598 | 931 | 319<br>(255, 383) | 1.52E-23 | 3.22E-23 |
| Dementia | 12,935 | 310 | 119<br>(89, 149) | 12,598 | 310 | 120<br>(84, 156) | 5.03E-11 | 8.69E-11 |
| Stroke | 12,935 | 318 | 106<br>(78, 134) | 12,598 | 318 | 90<br>(58, 121) | 3.39E-08 | 4.95E-08 |
| Lung Cancer | 12,935 | 130 | 83<br>(60, 107) | 12,598 | 130 | 78<br>(52, 104) | 2.85E-09 | 4.51E-09 |
| Prostate Cancer (Males Only) | 5,719 | 371 | 124<br>(67, 181) | 5,546 | 364 | 161<br>(94, 228) | 4.88E-06 | 6.18E-06 |
| Colorectal Cancer | 12,935 | 206 | 48<br>(26, 70) | 12,598 | 206 | 50<br>(24, 76) | 1.45E-04 | 1.72E-04 |
| Breast Cancer (Females Only) | 7,216 | 302 | 42<br>(-4, 89) | 7,052 | 302 | 32<br>(-24, 88) | 0.259 | 0.273 |
| Other Medical Conditions |  |  |  |  |  |  |  |  |
| Pneumonia | 12,678 | 712 | 355 (302, 408) | 12,351 | 712 | 354 (292, 416) | 3.55E-33 | 1.69E-32 |
| Chronic Kidney Disease | 12,793 | 557 | 272 (223, 321) | 12,457 | 557 | 281 (224, 338) | 1.52E-23 | 3.22E-23 |
| Delirium | 12,935 | 222 | 106 (78, 134) | 12,598 | 222 | 112 (79, 145) | 3.14E-11 | 5.96E-11 |
| Osteoarthritis | 11,378 | 1,723 | 222 (156, 288) | 11,084 | 1,723 | 191 (115, 267) | 1.12E-06 | 1.51E-06 |
| Osteoporosis | 12,666 | 434 | -22 (-49, 4) | 12,338 | 434 | 38 (6, 70) | 0.020 | 0.022 |
| Parkinson | 12,898 | 176 | 23 (4, 42) | 12,562 | 176 | 12 (-10, 35) | 0.285 | 0.285 |

<sup>1</sup>Change in cases per 0.1 unit decrease in HPS over 100,000 person-years of follow-up
<sup>2</sup>Fully adjusted covariates: age, sex, ethnicity, BMI, education, Townsend, smoking, hypertension, hypercholesterolemia, and UKB PPP consortium selection.

Lower HPS was significantly associated with the conditions in the healthspan definition (p_FDR-adj_<0.05), particularly mortality, diabetes, COPD, cancer, heart failure, and MI (**Table 1**). Among specific cancer diagnoses, a lower HPS was significantly associated with lung cancer, prostate cancer, and colorectal cancer (p_FDR-adj_<0.05) (**Table 1**). In addition, a lower HPS was associated with other medical conditions that were not included in the healthspan definition, such as pneumonia, chronic kidney disease, delirium, osteoarthritis, and osteoporosis (p_FDR-adj_<0.05) (**Table 1**).

For sensitivity analysis, we modeled HPS using four equal-interval groups and dichotomized HPS (“Low: HPS≤0.73 vs. High: HPS>0.73”). This threshold corresponds to the first quartile of HPS in the UKB PPP participants free from the conditions in the healthspan definition, effectively separating participants with and without these conditions at baseline (**Figure 1A**). Both analyses produced consistent results (**Supplementary Figures 5-8**). The Kaplan-Meier survival curves of the low and high HPS groups for the development of a first condition and individual conditions in the healthspan definition were clearly seprated in general, as presented in **Figure 2**.

**Figure 2.**
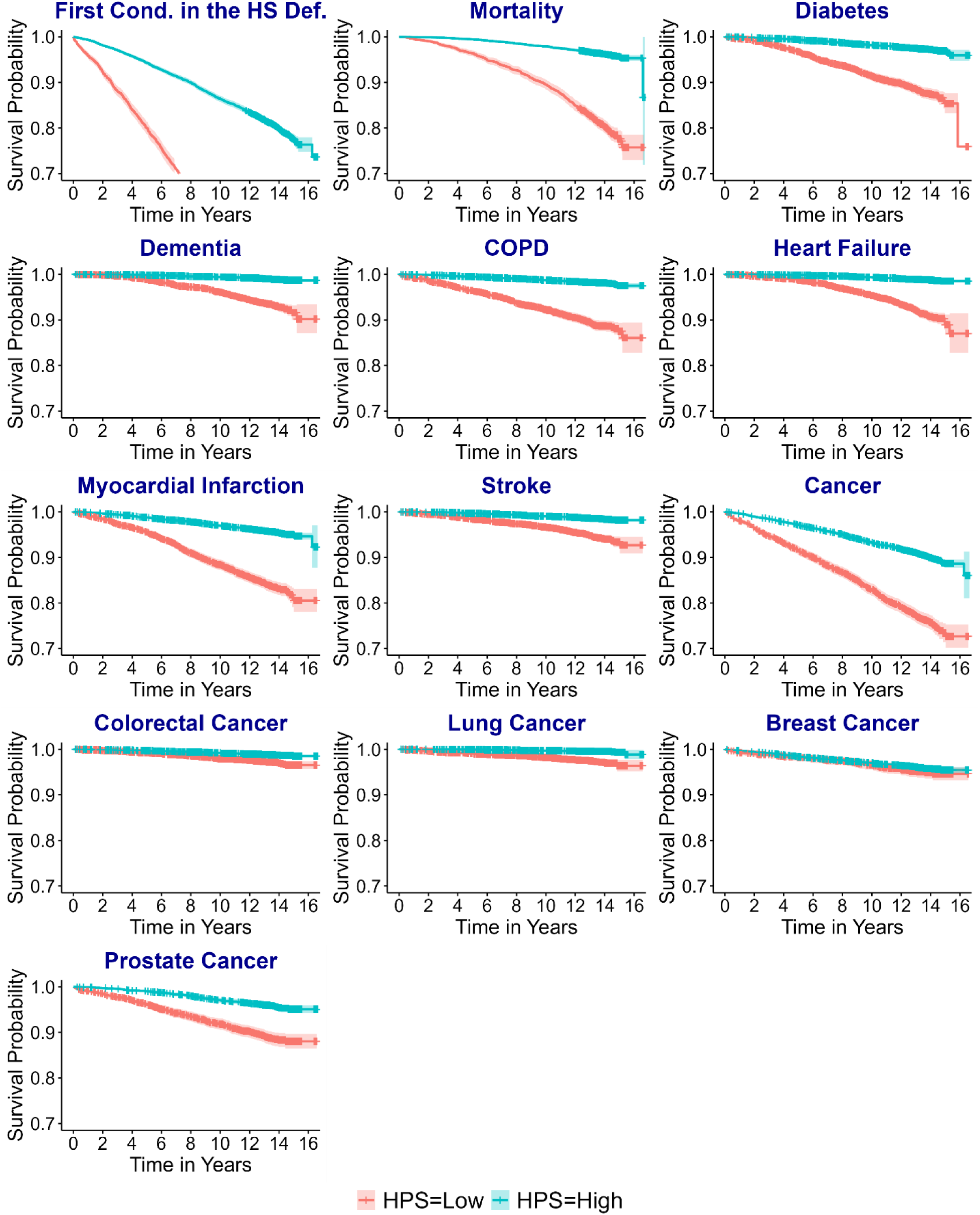
[UKB] Kaplan-Meier survival curves of the low (≤0.73) and high (>0.73) HPS groups for a first condition and individual conditions in the healthspan definition (n=12,935)

A low HPS score was associated with higher risks of a first condition in the healthspan definition, increased risk for the individual conditions, and other medical conditions (**Supplementary Figures 7 and 8**). Interestingly, the association between low HPS and healthspan was stronger in younger adults (<60 vs. ≥60 years), in previous or current smokers (vs. never smokers), and in obese participants (BMI≥30 vs. <30) (**Supplementary Figure 9**). Previous or current smokers also showed a stronger association with mortality (**Supplementary Figure 10**).

### HPS at baseline and a second condition in the healthspan definition or mortality during follow-up among UKB PPP participants with a specific healthspan condition only at baseline (cancer n=2,449, MI n=1,294, diabetes n=2,004, COPD n=1,412)

We further investigated how HPS at baseline is associated with the development of a second condition in the healthspan definition and mortality during follow-up, focusing on prevalent conditions at baseline: cancer, MI, diabetes, and COPD. HPS was strongly associated with a second healthspan condition and mortality during follow-up, regardless of the pre-existing condition (**Supplementary Figure 11**).

### Predictive power of HPS vs. systemic and organ-specific biological age measures (test sample, n=12,935)

PAC is a recently developed proteomics-based clock to predict mortality (15). Both HPS and PAC significantly predicted the conditions in the healthspan definition and other medical conditions (**Figure 3**). Compared to HPS and PAC, PhenoAge and ProtAge-EN were less predictive across conditions, except chronic kidney disease and osteoporosis, which were best predicted by PhenoAge and ProtAge-EN, respectively (**Figure 3**). We further compared HPS and PAC to organ-specific proteomic clocks to evaluate whether the latter are better predictors of organ-related conditions. Interestingly, HPS and PAC generally demonstrated similar or superior predictive performance compared to organ-specific clocks trained to predict chronological age or mortality (**Figure 4**).

**Figure 3.**
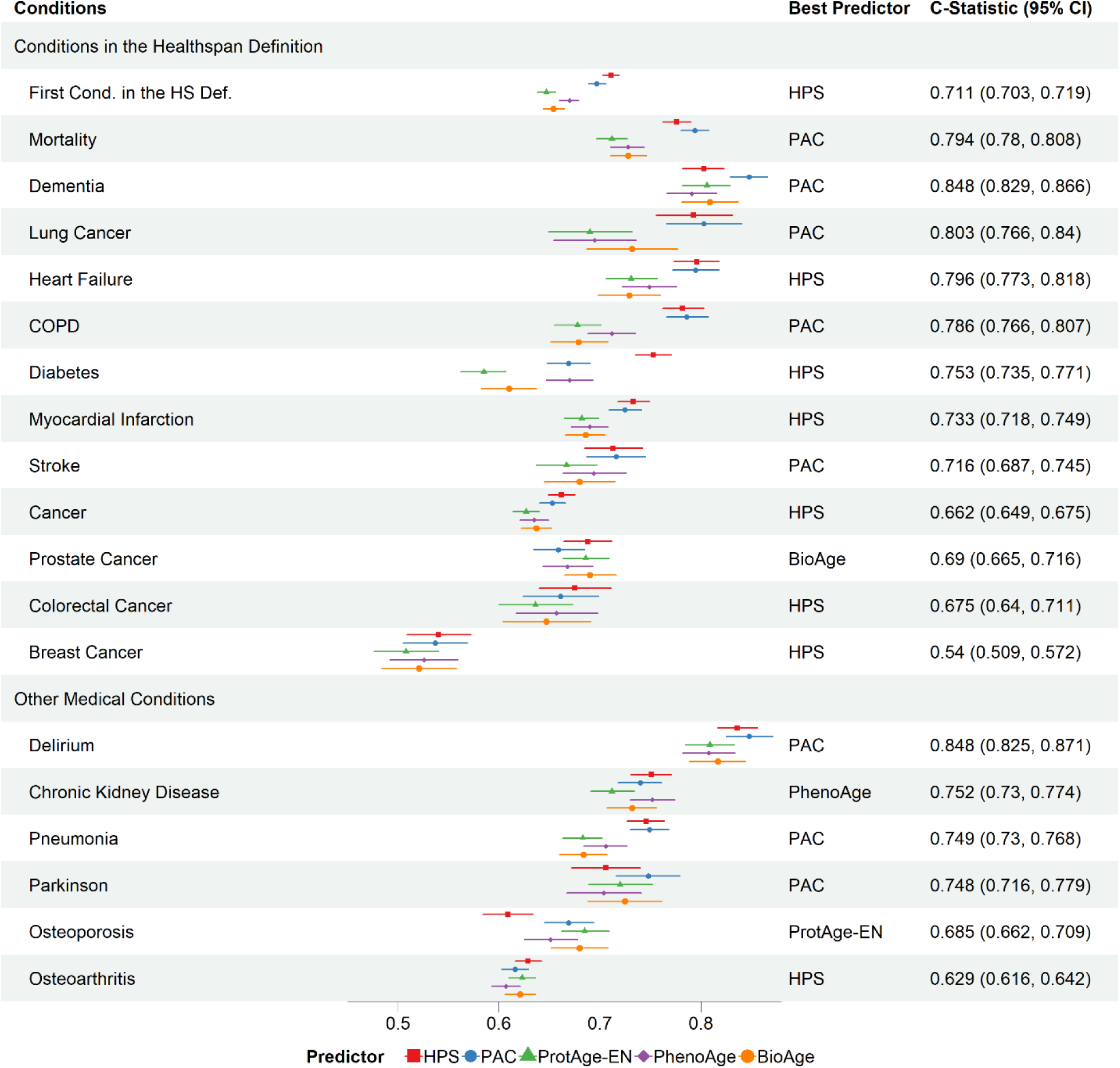
[UKB] Predictive power of healthspan proteomic score (HPS) versus proteomic aging clock (PAC), proteomic age (ProtAge), PhenoAge, and BioAge, assessed by C-statistics for time to a first condition and individual conditions in the healthspan definition (test sample, n=12,935).

**Figure 4.**
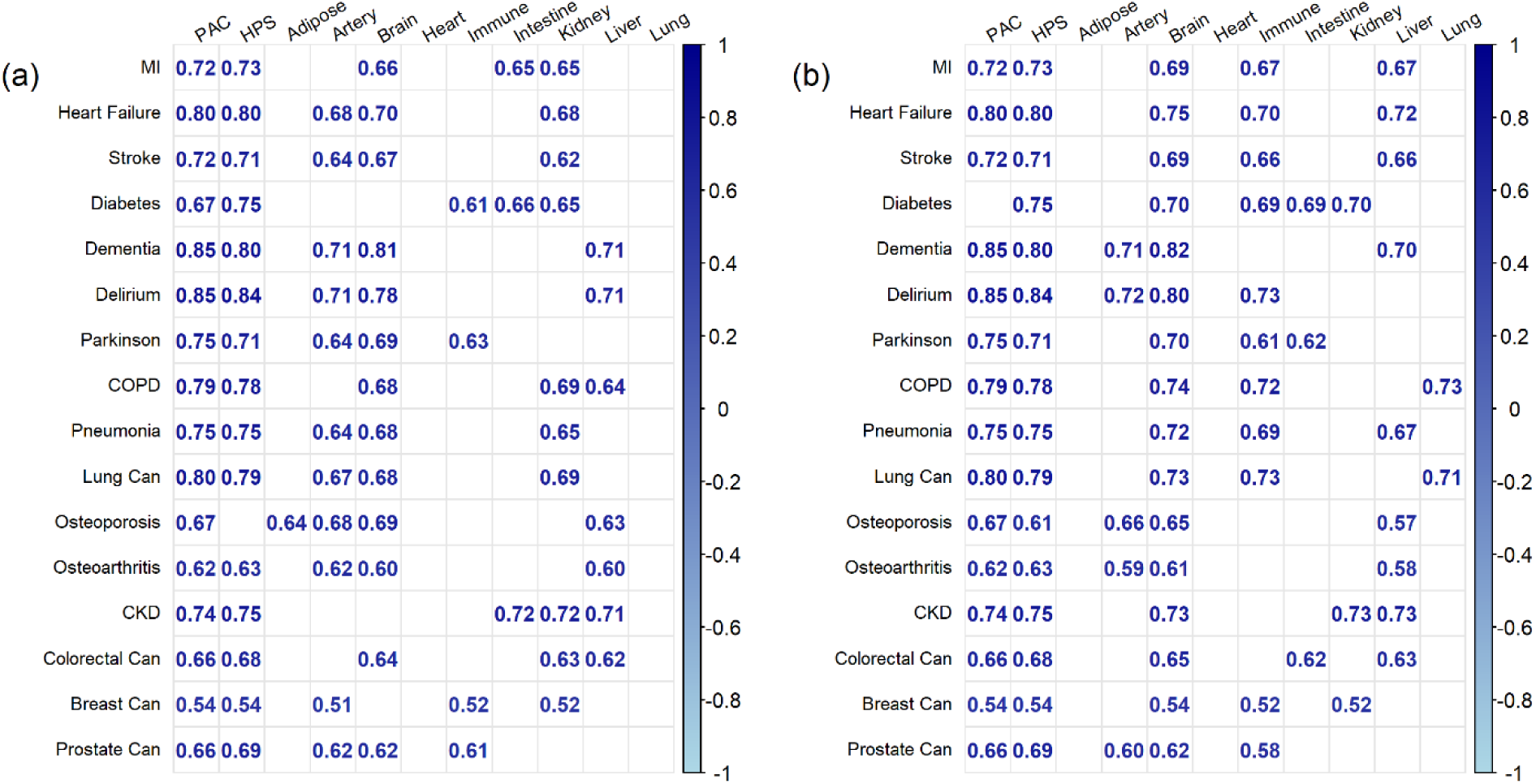
[UKB] Predictive power of healthspan proteomic score (HPS) and proteomic aging clock (PAC) versus organ-specific proteomic clocks trained to predict chronological age (a) or mortality (b), with top five C-statistics presented per condition

### Joint analysis of HPS and PAC groups at baseline and the risks of conditions in the healthspan definition, as well as other medical conditions during follow-up (test sample, n=12,935)

Despite their high predictive power for mortality and healthspan, HPS and PAC showed distinct predictive profiles for chronic diseases, suggesting that different biological processes underlie their measurements. We hypothesized that low HPS and high PAC significantly increased the risks of adverse health outcomes than low HPS or high PAC alone. We tested the synergistic effect of HPS and PAC on the associations with the conditions in the healthspan definition and other medical conditions. We categorized HPS and PAC at baseline using the cutoffs 0.73 and 58, respectively, corresponding to the first and third quartiles among UKB PPP participants without any condition in the healthspan definition at baseline. Either cutoff effectively separated participants with and without conditions included in the healthspan definition, as illustrated in **Figure 1A** and **Supplementary Figure 12**.

Among the participants in the test set with complete covariate data, 8,783 participants were in the high HPS & low PAC group (*better biological aging*), who were compared with 704 in the low HPS & low PAC group (*intermediate biological aging*), 762 in the high HPS & high PAC group (*intermediate biological aging*), and 2,349 in the low HPS & high PAC group (*worse biological aging*). A significant synergistic effect between low HPS and high PAC was found in the associations with the development of a first condition in the healthspan definition and mortality (FDR-adjusted interaction p=1.20×10^-5^ and p=0.002, respectively) (**Supplementary Figure 13**). Similar association patterns across the HPS and PAC groups were found with the conditions in the healthspan definition, such as heart failure, MI, stroke, and COPD (**Supplementary Figure 13**), as well as other medical conditions, including pneumonia and chronic kidney disease (**Supplementary Figure 14**).

### Gene set enrichment analysis of proteins associated with HPS (test sample, n=12,935)

Of 2,920 proteins analyzed, 1,398 showed significant associations (Bonferroni-corrected p<0.05) with low HPS (HPS≤0.73) after adjusting for covariates at baseline (age, sex, ethnicity, education, Townsend deprivation index, BMI, smoking status, hypertension, and hypercholesterolemia, and UKB PPP consortium selection) (**Supplementary Table 6**). Those significantly upregulated or downregulated proteins in the low HPS group are highlighted in **Figure 5A** (mean difference between the low and high HPS group after inverse normal transformation greater than 0.4 SD or smaller than 0.4 SD, chosen to label a maximal number of genes without losing clarity). The significant proteins (Bonferroni-corrected p<0.05) showed enrichment in 26 hallmark gene sets, including immune response, inflammation, cellular signaling, and metabolic regulation (**Figure 5B**).

**Figure 5.**
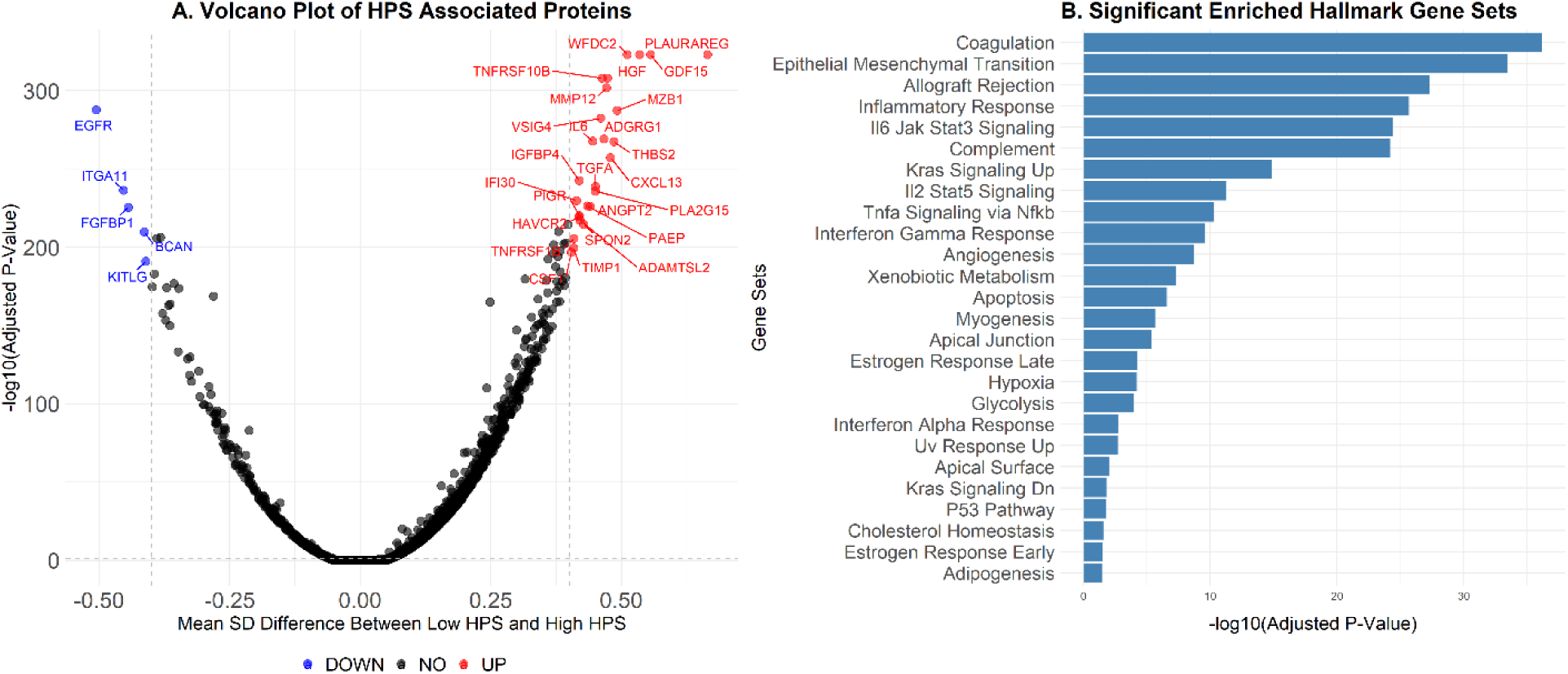
[UKB] HPS-associated proteins and enriched hallmark gene sets (test sample, n=12,935). **A.** Each point represents a protein, with the x-axis indicating the mean standard deviation (SD) difference in expression after inverse normal transformation (beta value) between the low and high HPS groups and the y-axis representing the statistical significance (-log10 adjusted p-value for multiple testing using Bonferroni correction). *Blue points* indicate significantly downregulated proteins in the low HPS group (beta < -0.4 and adjusted p-value < 0.05). *Red points* indicate significantly upregulated proteins in the low HPS group (beta > 0.4 and adjusted p-value < 0.05). *Black points* indicate proteins that are not significantly differentially expressed. **B**. The x-axis represents the significant gene sets, ordered by adjusted p-values for multiple testing, and the y-axis shows the -log10 of the adjusted p-value for each significant gene set (Bonferroni-corrected p-value <0.05).

### External replication using the EH-Epi study (n=401)

Participants in the EH-Epi study represent a subsample of the Finnish Twin Cohort (FTC) study. They were selected based on differing blood pressures between co-twins within the same family, as reported in a 2011 questionnaire from the Old FTC (25). This replication study included 401 twin individuals with available proteomic data. Among them, 41% were female and 47% had never smoked. The mean age was 62.3 years (range 56-70), with a mean BMI of 27.3 (SD = 4.9; range: 18-46). Of the 401 samples included, 379 had DNA methylation data for epigenetic age estimation, 399 had mortality follow-up, and 394 had blood metabolite data.

Of the 399 participants for whom mortality data was available, 13 deaths occurred over a median follow-up of 6.84 years, with seven attributed to cancer. Among these 13 deaths, 10 occurred in the low HPS group (HPS≤0.73) versus 3 in the high HPS group (HPS>0.73). The Kaplan-Meier survival curves of the low and high HPS groups for mortality are presented in **Figure 6**. The hazard ratio for a 0.1 unit decrease in HPS, adjusted for sex and chronological age, was 1.55 (95% CI 1.25 to 1.93, p<0.001), indicating that each 0.1 unit decrease in HPS increased the likelihood of mortality by 55%. The proportional hazards assumption of the Cox regression model was tested and not rejected (p=0.965).

**Figure 6.**
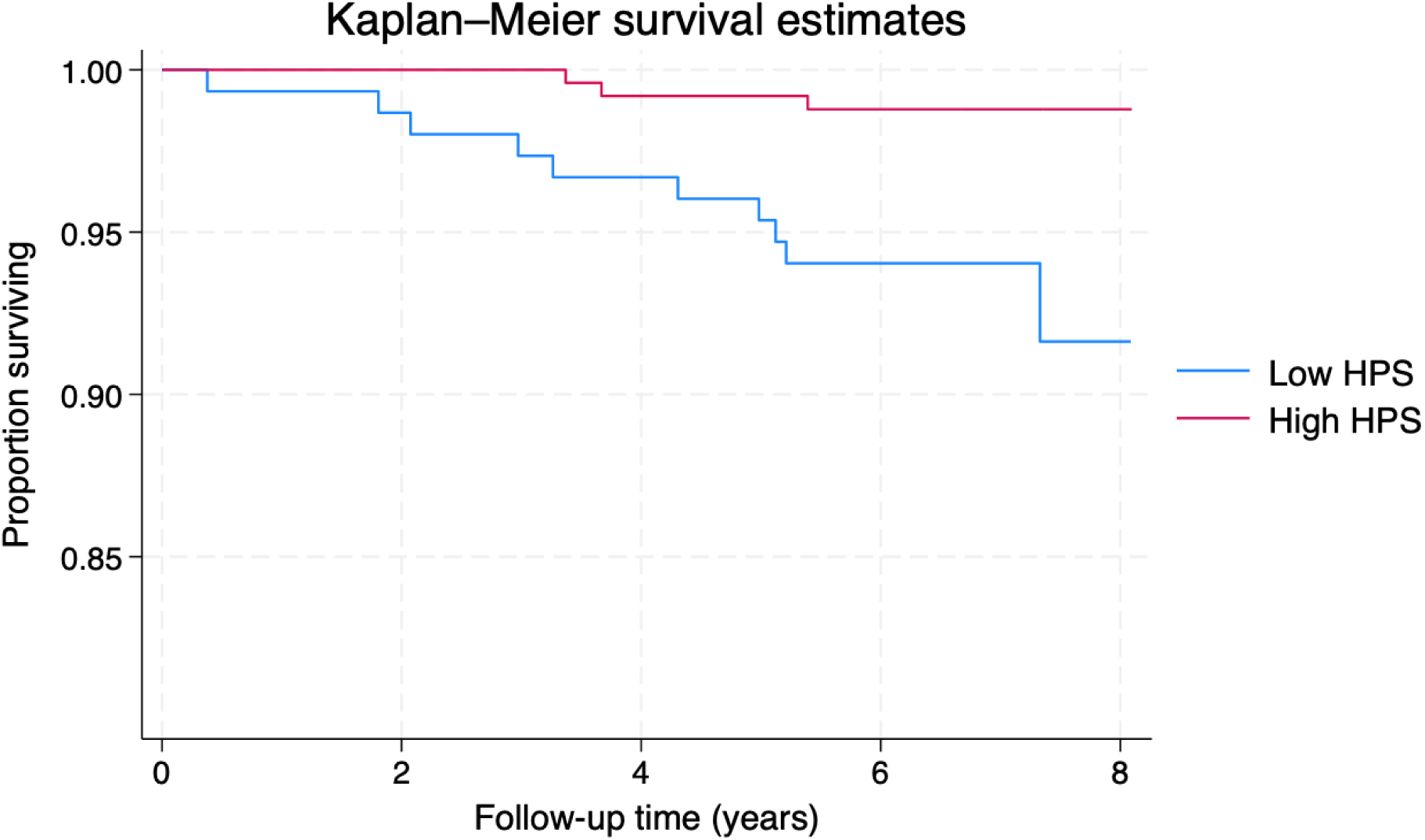
[EH-Epi] Kaplan-Meier survival curves of the low and high HPS groups for mortality (external validation sample, n=399)

HPS showed a weaker correlation with chronological age (Spearman correlation *r* = - 0.37) compared to the UKB data (*r*= -0.73). Six proteins required to calculate HPS had missing rates above 20%. For sensitivity analysis, we recalculated HPS excluding these proteins with high missing rates, while maintaining the same summed absolute weights. The results showed a minimal change in the correlation between HPS and chronological age (*r* = -0.40). Additionally, lower HPSs were more likely observed in males (p=3.3×10^-4^), older adults (≥60 years, p=7.9×10^-8^), current or former smokers (p=1.6×10^-4^), and obese individuals (BMI≥30, p=1.1×10^-11^) (**Supplementary Figure 15**).

HPS showed a strong correlation with PAC (*r*= -0.82) and low to modest correlations with epigenetic clocks, particularly GrimAge (*r*= -0.63) and GrimAge2 (*r*= -0.64) (**Supplementary Figure 16**). These correlations reduced after removing the effect of chronological age, although that between the residuals of HPS and DunedinPACE showed a slight increase (**Supplementary Figure 16**).

HPS performed similarly to DunedinPACE, both outperforming PAC and other epigenetic clocks in associations with hypertension (161 cases, **Supplementary Figure 17**) and blood metabolites (**Supplementary Figure 18**), after adjusting for sex and chronological age. These metabolites included albumin and three branched-chain amino acids (valine, leucine, isoleucine), recognized as markers of healthy aging (26, 27). HPS was not significantly associated with the cardiovascular outcome (14 cases with a history of coronary heart disease or stroke), in contrast to significant associations observed with Horvath, Hannum, and DNAm PhenoAge. Although trends were noted, none of the biological age measures were significantly associated with the pulmonary outcome (38 cases with a history of bronchial asthma, emphysema, chronic bronchitis, or COPD).

## Discussion

Unlike previous biological age predictors trained to predict chronological age, mortality (28) or organ/disease-specific predictors (21, 29–32), HPS was developed to evaluate the risk of a major chronic disease or mortality based on a healthspan definition. Therefore, HPS can be viewed as a systemic marker of biological aging. HPS outperformed chronological age and other biological age measures in predicting mortality and various chronic diseases. Additionally, it demonstrated better predictive performance for organ-related conditions than organ-specific proteomic clocks. Subgroup analyses revealed that individuals at higher risk for adverse health outcomes—such as males, older adults, current or former smokers, obese individuals, and those with hypertension or hypercholesterolemia—had lower HPS values, indicating a less healthy systemic biological status in these groups, pointing to a shared mechanism (i.e., decline in biological aging) that increase the risk of common adverse outcomes in these individuals. Additionally, we found that the biological processes associated with HPS align with the hallmarks of biological aging, particularly immune response, inflammation, cellular signaling, and metabolic regulation, providing a strong biological validation of HPS as an integrative measure of biological aging. HPS serves as a novel proteomic aging measure, complementing existing proteomic and epigenetic measures. Combining PAC and HPS strengthens the associations with mortality and multiple adverse health outcomes. Overall, our findings demonstrate that HPS is a valid biological aging measure, having strong clinical, predictive, and biological validity, making it a valuable tool for assessing healthspan in humans and potentially guiding the development of geroscience-based interventions in the future.

Several biological age measures have been developed to predict mortality and the onset of age-related diseases. These measures may reflect different aspects of biological aging due to data variations, study populations, and the methods used for their development (8, 33, 34). We compared HPS with several well-established biological age measures. HPS and PAC showed a strong correlation (*r* = -0.87); however, their correlation decreased significantly after adjusting for the effect of chronological age (*r*= -0.66). HPS, PAC, and other biological age measures are moderately correlated with one another, with chronological age being a significant contributor to these correlations. However, their residuals after removing the effect of chronological age show lower correlations between measures. This suggests that biological age acceleration based on different measures may reflect varying aspects of biological aging, which are captured uniquely by each biological age measure. These findings highlight the importance of leveraging multiple biological age measures to provide a more comprehensive assessment of biological aging in a given individual.

HPS may offer several advantages over other biological age measures, particularly in the context of geroscience-guided interventions. It was trained on relatively young individuals in a healthy cohort who had not developed chronic medical conditions at baseline and were followed until a first condition in the healthspan definition. HPS is a biological aging measure designed to capture the early manifestations of common chronic diseases, distinguishing it from existing epigenetic or proteomic clocks typically trained to predict chronological age or mortality. HPS strongly predicts mortality, and the onset of medical conditions in participants free from the conditions in the healthspan definition. Additionally, HPS predicts multimorbidities and mortality in those with a specific chronic medical condition. It generally demonstrates similar or superior predictive performance for organ-related conditions compared to organ-specific proteomic clocks. Multiple organs likely contribute to each condition, and predictors trained on shared outcomes such as mortality and healthspan benefit from the broader information captured across organ systems. While additional replication is necessary, HPS is effective in the EH-Epi study and holds promise for broader applications in healthier populations. By monitoring the effects of interventions on biological health, HPS can shorten trial lengths by serving as a surrogate measure for healthspan or chronic medical conditions. Therefore, incorporating HPS into clinical trials could enhance the evaluation of interventions to improve overall biological health and prevent chronic diseases.

The joint analysis of HPS and PAC revealed intriguing patterns of associations with mortality, and specific chronic medical conditions. Most individuals (70%) were classified in the better biological aging group (i.e., high HPS and low PAC). Meanwhile, 19% were in the worse biological aging group (i.e., low HPS and high PAC), and a small group, comprising 12% of the sample, were in the intermediate biological groups (i.e., those with low HPS and low PAC or high HPS and high PAC). As expected, individuals in the worse biological aging group showed the highest risk of adverse health outcomes. Those in the intermediate biological aging group also had an elevated risk, although the associations were not as strong as those in the worse biological aging group. Our findings imply that there is a group of “clinically” healthy individuals who already exhibit significant systemic biological changes, indicating a higher risk of disease development. This group may benefit the most from geroscience-guided interventions, particularly those in transitional stages of biological health (i.e., intermediate biological aging groups) who are at higher risks of diseases but could respond well to such interventions at an earlier stage in the pathophysiologic process.

Prior studies have evaluated proteomic markers associated with chronological age (35–37) and longevity, using various definitions of longevity, such as parental age at death above 95 years (38) or survival to 90 years (39). These studies identified robust proteomic signatures related to chronological age and longevity. Interestingly, a few markers are common across these studies, including ours, such as GDF-15, pleiotrophin, and chordin-like protein 1. Despite differences in proteomic platforms, proteome coverage, statistical modeling, and outcomes (e.g., chronological age, longevity, and healthspan), there appears to be a common proteomic signature that can capture multiple processes involved in healthy aging. Although still elusive, this common signature offers novel opportunities to develop interventions targeting multiple aging-related conditions.

Several limitations should be noted when interpreting our results. Disease diagnosis often occurs after the actual onset of pathological processes (diagnostic lag). As a result, some participants included in the HPS training set might have already experienced biological changes related to specific chronic medical conditions, which could influence the selection of proteins in the HPS model. To address this issue, we used multi-source data to determine the first diagnosis date, potentially reducing the impact of diagnostic lag. Moreover, since this diagnostic lag likely occurred randomly, it tends to drive associations toward the null. Consequently, our results may be more conservative than the actual associations (40).

Given the lack of consensus on the healthspan definition, predictors trained using different definitions may differ from HPS in assessing biological aging. We adopted a practical healthspan definition using electronic health records, which differs from the definition used by Walter et al. (41). They defined healthspan as the number of years from birth to the first occurrence of conditions in our healthspan definition plus hip fracture. A critique of both definitions is that, except for dementia, late-onset diseases are underrepresented, meaning those who remain disease-free may not necessarily be in good health (42). Including additional late-onset diseases is unlikely to significantly change our results, as the UKB is a relatively young cohort, and our goal was to train a predictor for early interventions.

Although our analyses focused on the conditions used to define healthspan, other diseases, such as chronic kidney disease, functional decline, and disability, could also significantly impact healthspan. Therefore, our results do not necessarily reflect a proteomic signature of the complete absence of all diseases. We have avoided terms like “disease-free” and “healthspan conditions” as our goal was not to define healthspan precisely but to use this definition as a framework for training a healthspan predictor in a healthier population. This approach aims to capture early biological changes of common chronic diseases, and our results support this objective. Thus, it is expected that HPS also shows strong associations with a broad range of outcomes beyond the specific conditions in the healthspan definition, after adjusting for chronological age and other covariates. While HPS was developed to test the geroscience hypothesis and serve as a systemic indicator of biological health, it often outperforms organ-specific proteomic clocks in predicting organ-related conditions. Finally, the UKB has limited racial and ethnic diversity, being predominantly composed of individuals of Caucasian ancestry. Likewise, the EH-Epi study consists of individuals of European ancestry with a homogeneous ethnic background. The external validation was primarily based on cross-sectional data. Although the association between HPS and mortality during follow-up was statistically significant, it was based on a small number of deaths and will require further validation in a larger cohort. Nonetheless, this head-to-head comparison is unique in that it incorporates both epigenetic and proteomic data, with the proteomic data generated using the same assay used in the UKB.

In conclusion, we have developed a novel healthspan proteomic score termed HPS, which demonstrated robust associations and predictions for mortality, and the onset of various diseases. The proteins associated with HPS were enriched in several processes related to the hallmarks of biological aging. HPS can be useful for gauging an individual’s biological health and monitoring the impact of geroscience-guided interventions, serving as a surrogate marker of healthspan. The field has recognized the importance of using multiple biological age measures, given the low to modest correlations between their residuals after controlling chronological age. HPS represents a valid proteomic measure. Our research shows that combining PAC and HPS significantly strengthens the associations with mortality and multiple adverse health outcomes, supporting the adoption of this integrative approach. Future studies are necessary to test whether HPS can be modulated by geroscience-guide interventions and serve as a surrogate marker for mortality and adverse health outcomes.

## Online Methods

### UK Biobank: HPS development and validation

#### Cohort description

The UKB is a volunteer community cohort in the United Kingdom that recruited over 500,000 participants aged 40 to 70 between 2006 and 2010 (43). During baseline assessments, data were collected on sociodemographic and lifestyle factors, environmental exposures, health and medical history, self-reported medications, cognitive function, physical activity, physical measurements, and biological samples for future assays. Over the years, various biological data have been generated, including plasma proteomic data from a nested cohort, as part of the UKB PPP (44). The baseline cohort participants have been followed up through linkages with electronic health records for disease diagnoses and deaths (45).

#### Proteomic data

The UKB PPP participants (initially n=54,219) were primarily a random sample (n=46,595, 85.9%) from the UKB baseline cohort. Additionally, the cohort included individuals selected by the UKB PPP consortium of 13 biopharmaceutical companies with specific disease or ancestry interests (n=6,376) and participants attending the COVID-19 repeat imaging study (n=1,268) (44). This study included proteomic data from 53,018 active UKB PPP participants after quality control. Protein concentrations of 2,923 proteins were measured in four panels (cardiometabolic, inflammation, neurology, and oncology) using the Olink Explore 3072 platform and normalized through a two-step process involving within-batch and across-batch intensity normalization (46). These panels are pre-designed to target specific sets of proteins based on biological relevance, assay performance, and prior applications in biomedical research to maximize their utility for studying complex traits and diseases. The normalized protein expression (NPX) data were used throughout the present project. Three proteins, GLIPR1 (99.7%), NPM1 (74.0%), and PCOLCE (63.6%), were excluded due to high missing rates. The median individual missing rate was 0.5% (25^th^ percentile 0.1% and 75^th^ percentile 7.5%). However,7.2% of the included samples only had complete proteomic data, necessary for the HPS development methods. To address this issue, we imputed the missing proteomic data using the *k*-nearest neighbors approach (*k*=10) with the R package ‘multiUS’ (47).

#### Healthspan definition

Healthspan was defined as the number of years from birth free of cancer (excluding non-melanoma skin cancer), diabetes (including type I diabetes, type II diabetes, and malnutrition-related diabetes), heart failure, MI, stroke, COPD, dementia, or death, in line with previous studies (16, 17). The UKB acquired the date of death data through linkages with national death registries and derived the first diagnosis dates for the conditions in the healthspan definition and other medical conditions based on ICD-10 codes (**Supplementary Table 2**), which were used to link multi-source data, including primary care records, hospital inpatient data, cancer and death registries records, and self-reported medical condition codes.

#### HPS development

Participants without any condition in the healthspan definition at baseline were randomly split into a training set and a test set in a seven-to-three ratio (**Supplementary Figure 1**). The training set data, including chronological age and NPX of 2920 proteins, was related to time from baseline to a first condition in the healthspan definition (censored at death or last follow-up of hospital inpatient data [main source], whichever occurred first) using a LASSO Cox regression model. The death censoring date was 2022/11/30. The inpatient censoring dates were 2022/11/30 for participants attending baseline assessment centers in England, 2021/7/31 for participants in Scotland, and 2018/2/28 for those in Wales. Proteins with non-zero regression coefficients in the fitted model were carried forward to fit a Gompertz regression model. Based on the cumulative density probability of this model, the risk of developing a first condition in the healthspan definition within 10 years from baseline can be derived (*r*), with 1-*r* representing the likelihood of remaining free from the conditions in the healthspan definition, referred to as the healthspan proteomic score (HPS).

#### HPS validation

We tested the validity of HPS using participants in the test set by examining:

- Spearman correlations of HPS with chronological age, biological age measures, as well as aging traits at baseline, where the biological age measures of PAC (15), ProtAge-EN (21) (protein-predicted age trained using the elastic net method (21), distinguished from that developed by Argentieri et al. (48), trained using the light gradient boosting machine method), PhenoAge (11), BioAge (22), and a 49-item frailty index (23) were derived using data fields listed in **Supplementary Table 2.**
- Spearman correlations between the residuals of HPS, PAC, ProtAge-EN, PhenoAge, and BioAge after removing the effect of chronological age in linear regression models, and baseline aging traits.
- Associations of HPS at baseline with disease risk factors at baseline (age, sex, smoking, obesity, hypertension, and hypercholesterolemia), mortality, and medical conditions during follow-up (detailed in **Supplementary Table 2**).
- Associations of HPS at baseline with a first condition in the healthspan definition and mortality during follow-up in subgroups by age (≥60, <60), sex (male, female), smoking status (previous or current smokers, never smokers), BMI (≥30, <30), hypertension (yes/no), and hypercholesterolemia (yes/no).
- Predictions for the conditions in the healthspan definition and other medical conditions against systemic (ProtAge-EN, PAC, PhenoAge, and BioAge) and organ-specific proteomic clocks (adipose, artery, brain, heart, immune, intestine, kidney, liver, lung, and muscle) trained to predict chronological age or mortality by Goeminne et al. (21).
- The synergistic effect of low HPS (HPS≤0.73) and high PAC (PAC>58) on the associations with the conditions in the healthspan definition and other medical conditions. during follow-up.
- The validity of HPS in those with a specific condition in the healthspan definition at baseline for associations with a second condition and mortality.

### Statistical Methods

Between-group comparisons of HPS based on a disease risk factor were conducted using Wilcoxon rank-sum tests. Association analyses used Aalen’s additive hazard models (49) adjusting for baseline covariates (chronological age, sex, ethnicity, education, Townsend deprivation index, BMI, smoking status, hypertension, hypercholesterolemia, and UKB PPP consortium selection, **Supplementary Table 2**), which facilitated testing for an additive interaction effect between HPS and PAC (50). Throughout the association analysis, p-values were adjusted for multiple testing using the Benjamini-Hochberg false discovery rate method (51). Harrell’s C statistic (52) under a Cox regression model was used to assess the comparative predictive accuracy for mortality, and incident medical conditions during follow-up.

To understand the measurement of HPS, we conducted a gene set enrichment analysis for proteins associated with low HPS (HPS≤0.73) after adjusting for baseline covariates. Prior to the association analysis, the inverse normal transformation was applied to each protein to normalize the distribution and unify the scale into z-scores. Proteins significant at the Bonferroni-corrected level were entered into the gene set analysis implemented in the Functional Mapping and Annotation of Genome-Wide Association Studied (FUMA version 1.5.2). Genes associated with HPS were compared with the background genes (20,260 protein-coding genes) for the presence in a hallmark gene set using a hypergeometric test. Enriched hallmark gene sets with at least five genes overlapped with the input genes were identified at the Bonferroni-corrected level of 5% (50 hallmark gene sets in total).

### The Essential Hypertension Epigenetics (EH-Epi) study: external validation of HPS Cohort description

The external replication analyses were performed in an independent sample of twins from the Finnish Twin Cohort (FTC) study who were intensively studied and phenotyped as part of a substudy: the Essential Hypertension Epigenetics (EH-Epi) study. Twins were initially identified from the older FTC study questionnaire sent in 2011 (25). Liked-sex twin pairs in which the twins differed in blood pressure were invited to participate in the EH-Epi study. The twins provided fasting blood samples during in-person visits from 2013 to 2015, during which additional measures were collected (53). A variety of omics data, including the proteomics and metabolomics, and DNA methylation data used in the current study, were generated from the blood samples.

### Proteomic data

Proteomic data (Olink Proteomics AB, Uppsala, Sweden; Olink Explore 3072) were originally generated in 415 EH-Epi twins from plasma samples, as described in detail elsewhere (54). The data were quality controlled according to Olink’s internal quality control criteria using the R package ‘OlinkAnalyze’ version 3.4.1, which resulted in the rejection of a few outlier samples. Using the normalized protein expression (NPX) data, we calculated PAC and HPS estimates in 401 twins with relevant summary statistics.

### DNA methylation data

DNA methylation levels were quantified using the Infinium Illumina HumanMethylation450K array and preprocessed using the R package ‘meffil’ (55), as described in detail elsewhere (56). In the current study, we used six previously generated epigenetic age estimates available for 379 of 401 Finnish twins. Horvath (57), Hannum (58), DNAm PhenoAge (11), and GrimAge (13) estimates were calculated using their PC score versions (59), as described elsewhere (60). We also generated DunedinPACE estimates (61), which represent individuals’ pace of aging based on baseline methylation levels, and the latest versions of GrimAge, GrimAge2 (62).

### HPS validation

We tested the external validity of HPS in the EH-Epi study by examining:

- Association between HPS and mortality during follow-up, up to the year 2020, which was within 7 years of the blood sample collection. Date of death was provided by the Finnish population information system (https://dvv.fi).
- Spearman correlations of HPS with chronological age, PAC, ProtAge-EN, and epigenetic clocks.
- Spearman correlations between the residuals of HPS, PAC, and epigenetic clocks after removing the effect of chronological age in linear regression models.
- Associations of HPS with disease risk factors (age, sex, smoking, and obesity) and age-related outcomes at the time of blood sampling.

◦ Hypertension status defined as a systolic blood pressure (SBP)>140 mm Hg and a diastolic blood pressure (DBP)>90 mm Hg, after being corrected for medication use.
◦ Two binary outcomes based on the old FTC 2011 questionnaire. 1) Cardiovascular Outcome: Cases were twins who reported having been diagnosed with coronary heart disease or stroke; 2) Pulmonary Outcome: Cases were twins who reported having been diagnosed with bronchial asthma, emphysema, chronic bronchitis, or COPD.
◦ Finally, we used blood metabolomic data to examine albumin levels, since albumin is a known marker of aging, as well as the three branched-chain amino acids (valine, leucine, isoleucine), which have been linked to aging and age-related diseases such as type 2 diabetes. Metabolites were quantified using Nightingale nuclear magnetic resonance (NMR) technology, quality controlled as described elsewhere (63).

### Statistical Methods

Kaplan-Meier survival curves were plotted to compare mortality between the low (≤0.73) and high (>0.73) HPS groups. The association between HPS (as a continuous variable) and mortality was evaluated using a Cox proportional hazards model. Participants were entered into the analysis at the time of blood sampling and were followed until death or end of follow-up on December 31, 2020. Family relatedness was accounted for in the survival analysis. Both sex and chronological age at the time of blood sampling were included as covariates. The proportional hazards assumption of the Cox regression model was tested. The hazard ratio for a 0.1 decrease in HPS, 95% confidence interval, and p-value were reported.

Between-group comparisons of HPS based on disease risk factors were conducted using generalized estimating equation (GEE) models to account for familial relatedness. GEE models also were used to link HPS with each age-related outcome at baseline, with adjustments for chronological age and sex. PAC, ProtAge-EN, and epigenetic clocks were similarly analyzed for comparison. For the above analyses, biological age estimates were transformed using the rank-based inverse normal transformation to unify the scales and correct distributional skewness.

## Author Contributions

CLK and BSD designed the study. CLK and PL processed and analyzed the UKB data. CLK and BSD drafted the initial manuscript, which included contributions from PL. They also coordinated with GD, EV, MO, and JK to replicate the UKB findings using the EH-Epi study data. MO provided processed DNA methylation data and associated epigenetic age estimates used in the current study. All the authors reviewed and approved the final version.

## Data Availability Statements

Data access to the UK Biobank is granted upon application. The EH-Epi study data used in the analysis is available through the Biobank of the Finnish Institute for Health and Welfare (https://thl.fi/en/web/thl-biobank/forresearchers). It is available to researchers after written application and following the relevant Finnish legislation. The R code for computing PAC, HPS, and organ-specific proteomic clocks can be obtained from the GitHub repositories at https://github.com/kuo-lab-uchc.

## Acknowledgments

Access to UK Biobank data was granted under application no. 92647 “Research to Inform the Field of Precision Gerontology” (PI: Richard H. Fortinsky). This research used data assets made available by National Safe Haven as part of the Data and Connectivity National Core Study, led by Health Data Research UK in partnership with the Office for National Statistics and funded by UK Research and Innovation (research which commenced between 1 October 2020–31 March 2021 grant ref MC_PC_20029; 1 April 2021–30 September 2022 grant ref MC_PC_20058). This research also used data provided by patients and collected by the NHS as part of their care and support. Copyright © (year), NHS England. Re-used with the permission of the NHS England [and/or UK Biobank]. All rights reserved.

