## Supplementary Figures for "A proteomic signature of healthspan"

**Figure S1** [UKB] Sample selection for training (n=30,184) and testing (n=12,935) the healthspan proteomic score (HPS).

**Figure S2** [UKB] Baseline healthspan proteomic score (HPS) distributions (test sample, n=12,935) comparing (a) males vs. females, (b) older adults vs. younger adults, (c) previous or current smokers vs. non-smokers, (d) obese vs. non-obese participants, (e) hypertension vs. non-hypertension participants, (f) hypercholesterolemia vs. non-hypercholesterolemia participants.

**Figure S3** [UKB] Healthspan proteomic score (HPS) Spearman correlations with chronological age, other biological age measures, physiological, cognitive, and frailty measures at baseline (test sample, n=12,935).

**Figure S4** [UKB] Spearman correlations between residuals of healthspan proteomic score (HPSRes), proteomic aging clock (PACRes), ProtAge-EN (ProtAge-ENRes), PhenoAge (PhenoAgeRes), and BioAge (BioAgeRes) after removing the effect of chronological age using linear regression models, as well as physiological, cognitive, and frailty measures at baseline (test sample, n=12,935)

**Figure S5** [UKB] Risk differences comparing healthspan proteomic score (HPS) of (0.5, 0.75], (0.25, 0.5], or (0, 0.25) to HPS (0.75, 1] at baseline for developing a first condition or a specific condition in the healthspan definition (test sample, n=12,935).

**Figure S6** [UKB] Risk differences comparing healthspan proteomic score (HPS) of (0.5, 0.75], (0.25, 0.5], or (0, 0.25) to HPS (0.75, 1] at baseline for developing incident medical conditions not included in the healthspan definition (test sample, n=12,935).

**Figure S7** [UKB] Risk differences comparing low healthspan proteomic score (HPS≤0.73) to high HPS (>0.73) for developing a first condition or a specific condition in the healthspan definition (test sample, n=12,935).

**Figure S8** [UKB] Risk differences comparing low healthspan proteomic score (HPS≤0.73) to high HPS (>0.73) for developing incident medical conditions not in the definition of healthspan (test sample, n=12,935).

**Figure S9** [UKB] Risk differences comparing low healthspan proteomic score (HPS≤0.73) to high HPS (>0.73) for developing a first condition in the healthspan definition in subgroups (test sample, n=12,935).

**Figure S10** [UKB] Risk differences comparing low healthspan proteomic score (HPS≤0.73) to high HPS (>0.73) for mortality in subgroups (test sample, n=12,935).

**Figure S11** [UKB] Risk differences comparing healthspan proteomic score (HPS) of (0.5, 0.75], (0.25, 0.5], or (0, 0.25) to HPS (0.75, 1] at baseline for a second condition (a), or mortality (b), in participants initially presenting with a condition in the healthspan definition at baseline (cancer n=2,449, MI n=1,294, diabetes n=2,004, COPD n=1,412).

**Figure S12** [UKB] PAC proteomic age distributions in UKB PPP participants (n=53,018), separated by the number of conditions in the healthspan definition at baseline (red dashed line at 58, corresponding to the third quartile)

**Figure S13** [UKB] Risk differences comparing more adverse HPS and PAC groups to high HPS and low PAC group for a first condition and individual conditions in the healthspan definition (test sample, n=12,935). Fully adjusted covariates: age, sex, ethnicity, BMI, education, Townsend, smoking, hypertension, hypercholesterolemia, and UKB PPP consortium selection.

**Figure S14** [UKB] Risk differences comparing more adverse HPS and PAC groups to high HPS and low PAC group for incident medical conditions not in the healthspan definition (test sample, n=12,935).

**Figure S15** [EH-Epi] Baseline healthspan proteomic score (HPS) distributions (n=401) comparing (a) males vs. females, (b) older adults vs. younger adults, (c) previous or current smokers vs. non-smokers, (d) obese vs. non-obese participants.

**Figure S16** [EH-Epi] (a) Spearman correlations between HPS (n=401), PAC (n=401), and epigenetic clocks (n=399) and (b) correlations between their residuals after removing the effect of chronological age using linear regression models.

**Figure S17** [EH-Epi] Associations of HPS (n=401), PAC (n=401), and epigenetic clocks (n=379) with hypertension, cardiovascular, and pulmonary outcomes at the time of blood sampling, adjusted for sex and chronological age.

**Figure S18 [EH-Epi] Associations of HPS, PAC, and epigenetic clocks with blood metabolites (n=394) at the time of blood sampling, adjusted for sex and chronological age.**

UKB PPP active participants

Diagnosed with any condition in the healthspan definition before or at baseline

Free from the conditions in the healthspan definition

HPS training set

Remaining free from the conditions in the healthspan definition

Diagnosed with any condition in the healthspan definition during follow-up

HPS test set

Remaining free from the conditions in the healthspan definition

Diagnosed with any condition in the healthspan definition during follow-up

**Figure S1** [UKB] Sample selection for training (n=30,184) and testing (n=12,935) the healthspan proteomic score (HPS)

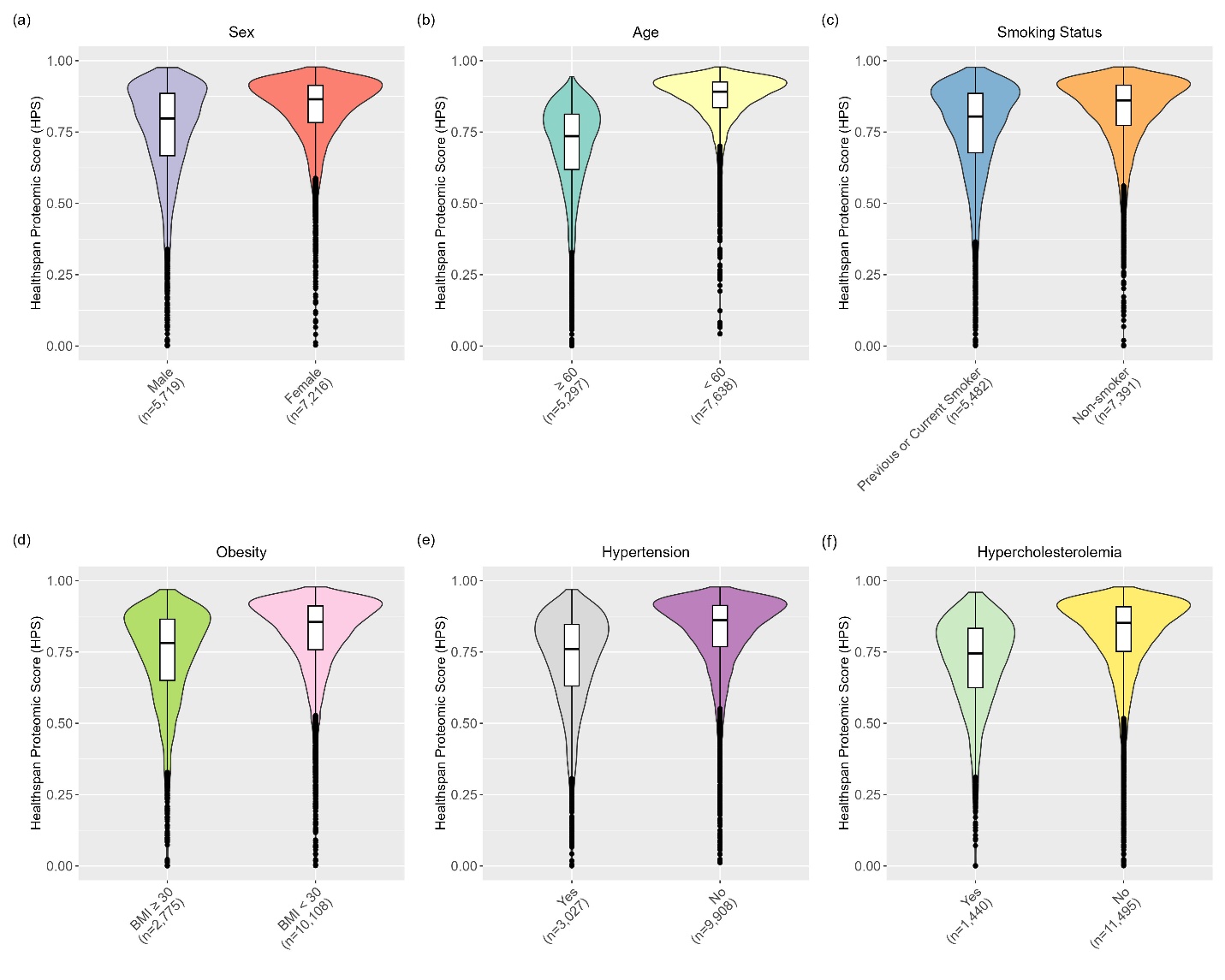

**Figure S2** [UKB] Baseline healthspan proteomic score (HPS) distributions (test sample, n=12,935) comparing (a) males vs. females, (b) older adults vs. younger adults, (c) previous or current smokers vs. non-smokers, (d) obese vs. non-obese participants, (e) hypertension vs. non-hypertension participants, (f) hypercholesterolemia vs. non-hypercholesterolemia participants.

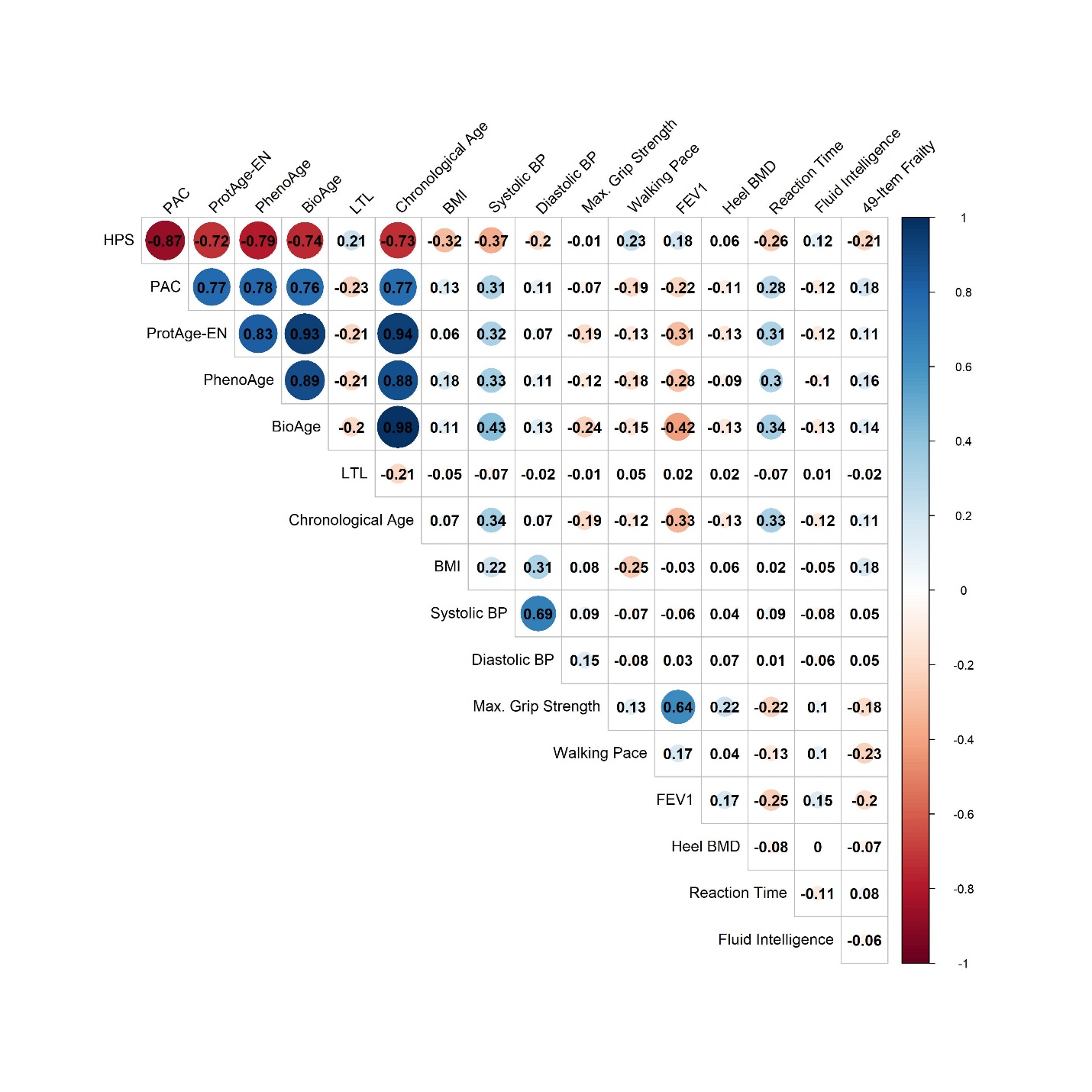

**Figure S3** [UKB] Healthspan proteomic score (HPS) Spearman correlations with chronological age, other biological age measures, physiological, cognitive, and frailty measures at baseline (test sample, n=12,935).

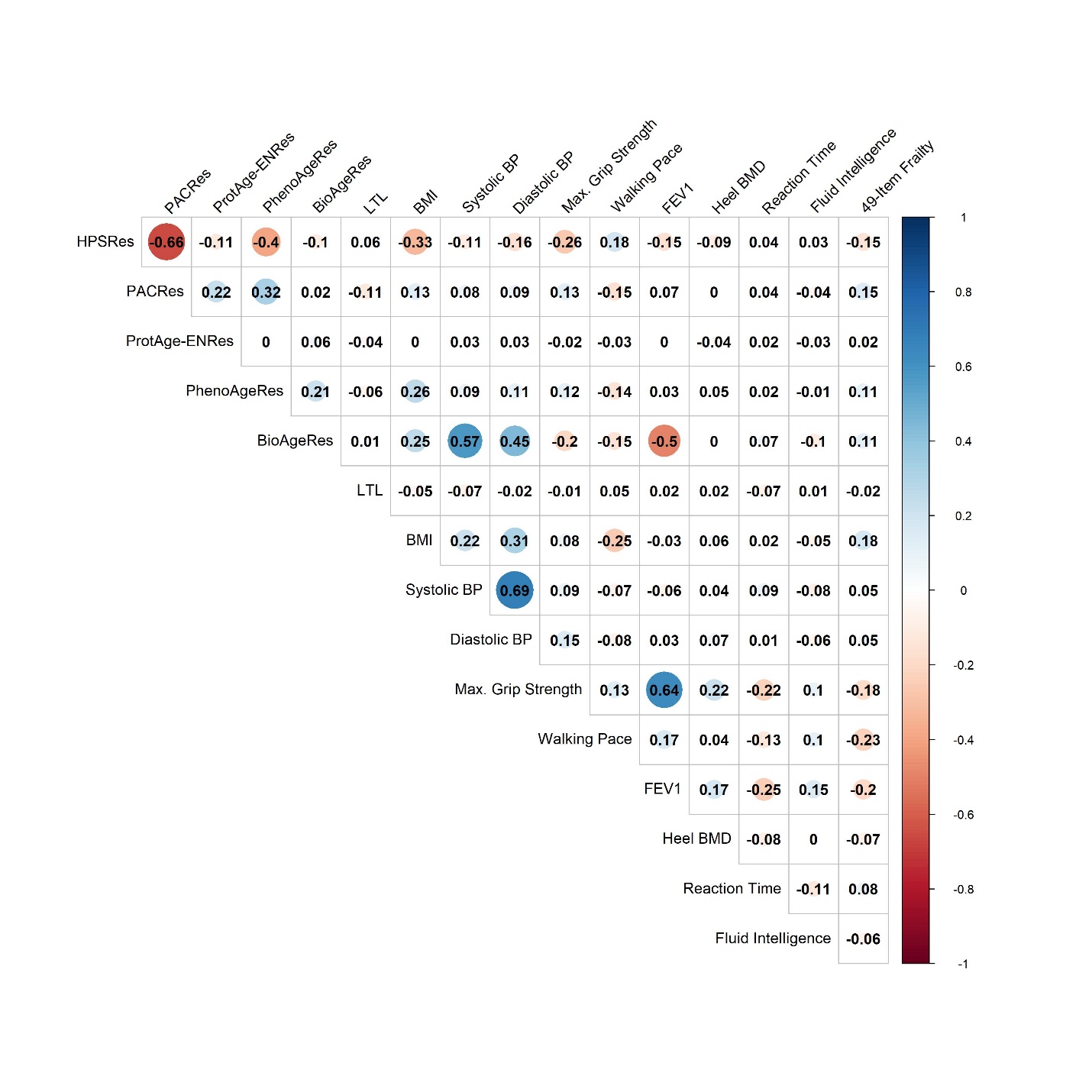

**Figure S4** [UKB] Spearman correlations between residuals of healthspan proteomic score (HRSRes), proteomic aging clock (PACRes), ProtAge-EN (ProtAge-ENRes), PhenoAge (PhenoAgeRes), and BioAge (BioAgeRes) after removing the effect of chronological age using linear regression models, as well as physiological, cognitive, and frailty measures at baseline (test sample, n=12,935).

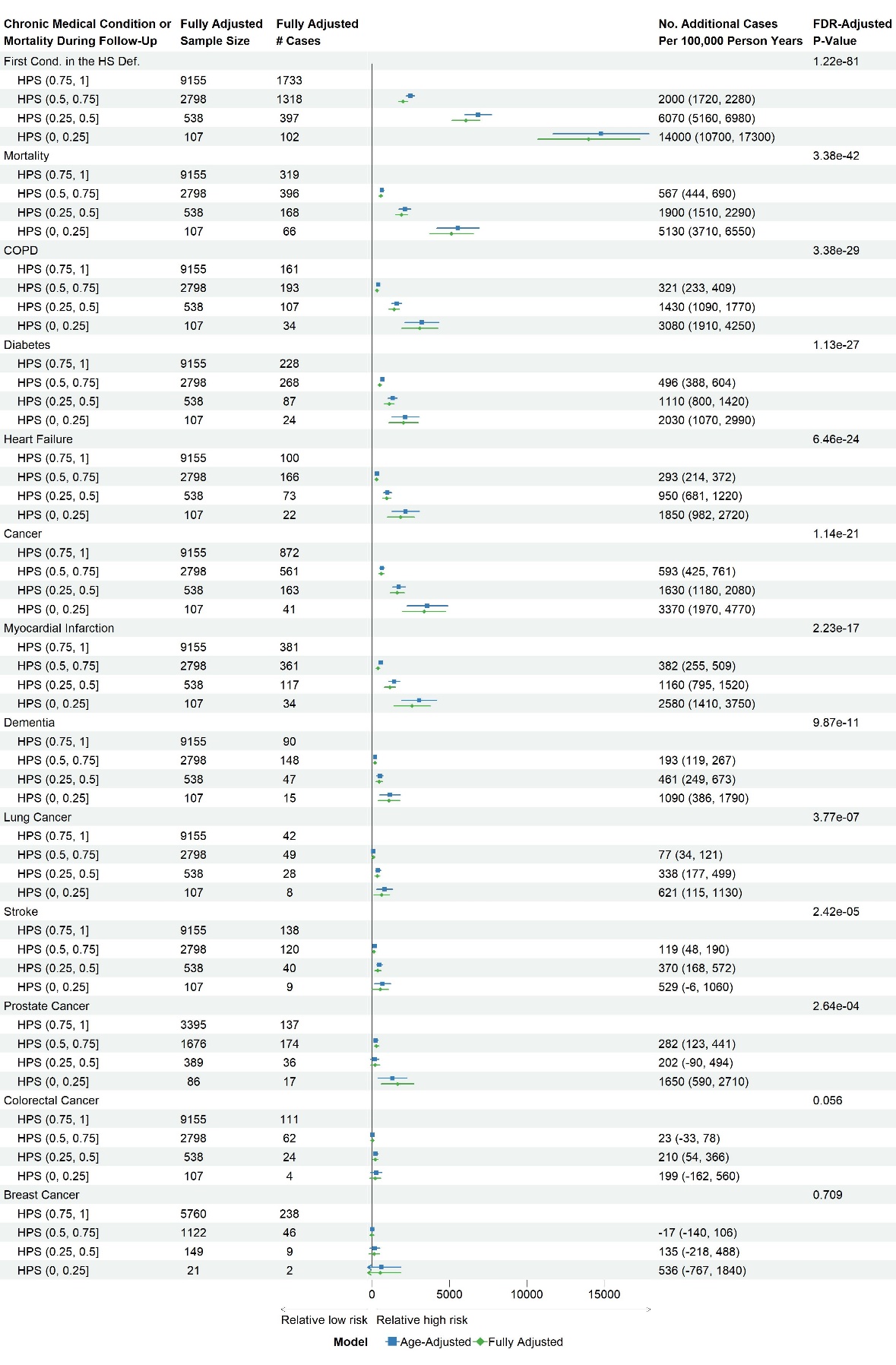

**Figure S5** [UKB] Risk differences comparing healthspan proteomic score (HPS) of (0.5, 0.75], (0.25, 0.5], or (0, 0.25) to HPS (0.75, 1] at baseline for developing a first condition or a specific condition in the healthspan definition (test sample, n=12,935).

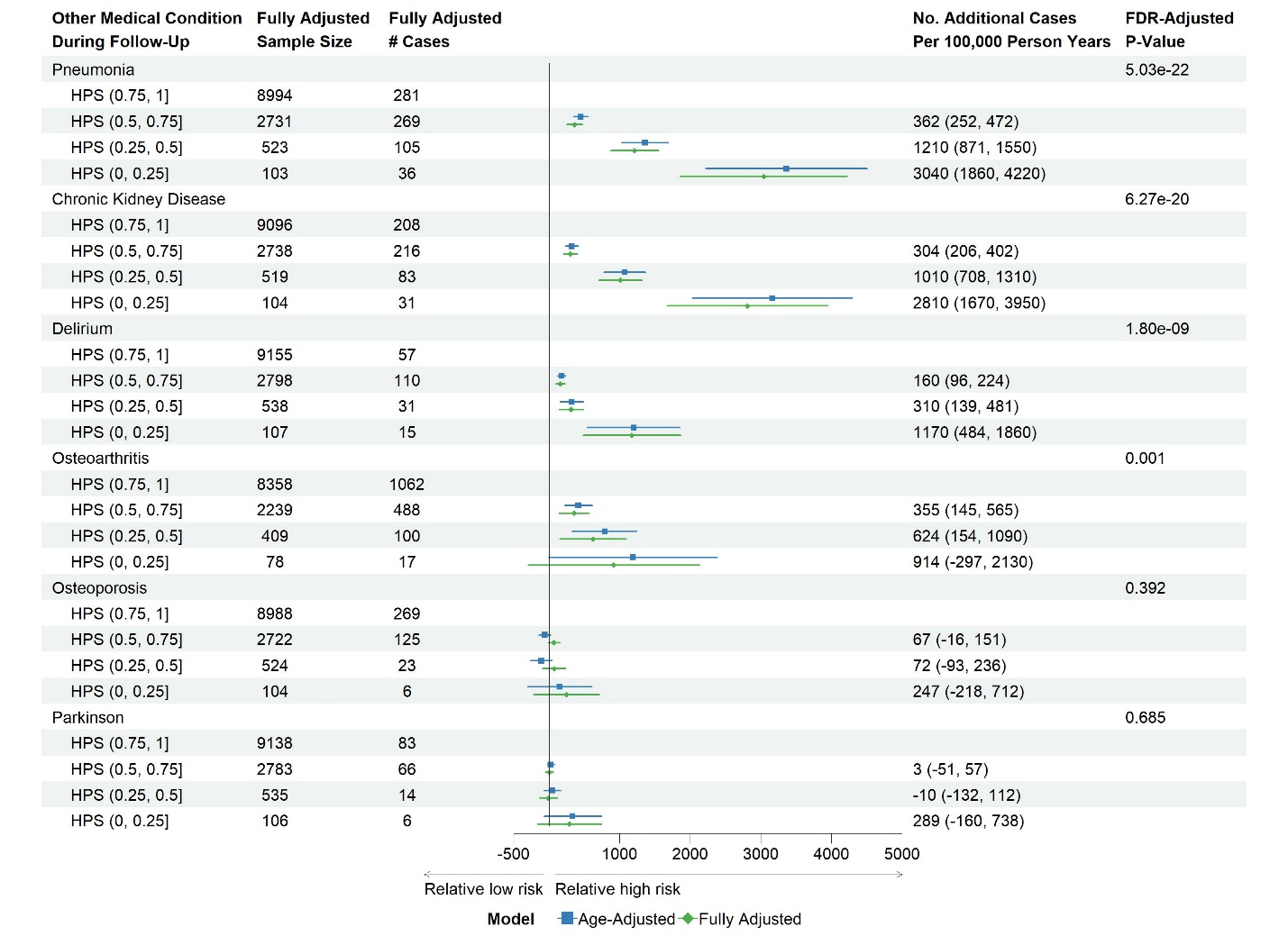

**Figure S6** [UKB] Risk differences comparing healthspan proteomic score (HPS) of (0.5, 0.75], (0.25, 0.5], or (0, 0.25) to HPS (0.75, 1] at baseline for developing incident medical conditions not included in the healthspan definition (test sample, n=12,935). Fully adjusted covariates: age, sex, ethnicity, BMI, education, Townsend deprivation index, smoking status, hypertension, hypercholesterolemia, and UKB PPP consortium selection.

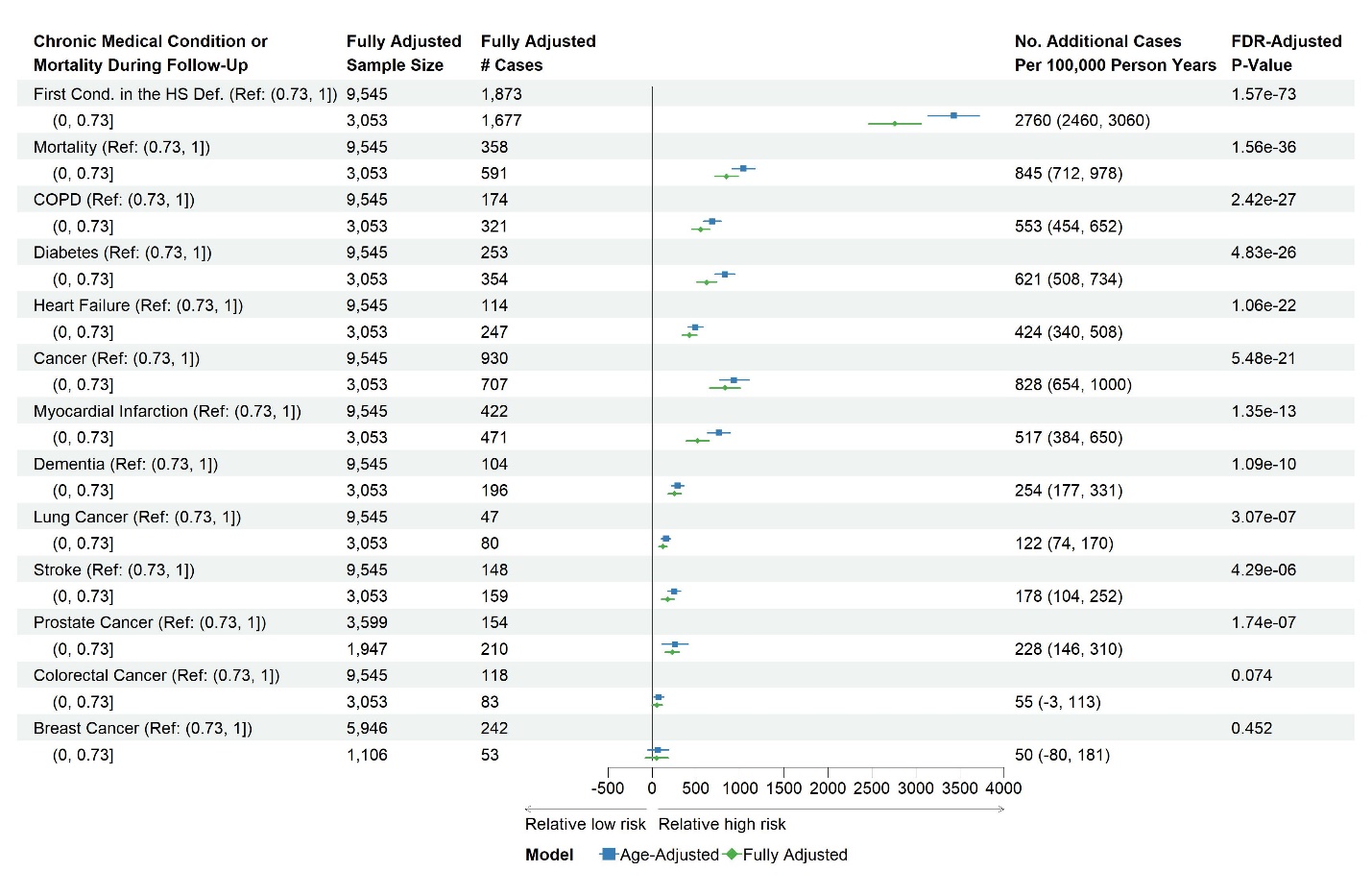

**Figure S7** [UKB] Risk differences comparing low healthspan proteomic score (HPS≤0.73) to high HPS (>0.73) for developing a first condition or a specific condition in the healthspan definition (test sample, n=12,935). Fully adjusted covariates: age, sex, ethnicity, BMI, education, Townsend, smoking status, hypertension, hypercholesterolemia, and UKB PPP consortium selection.

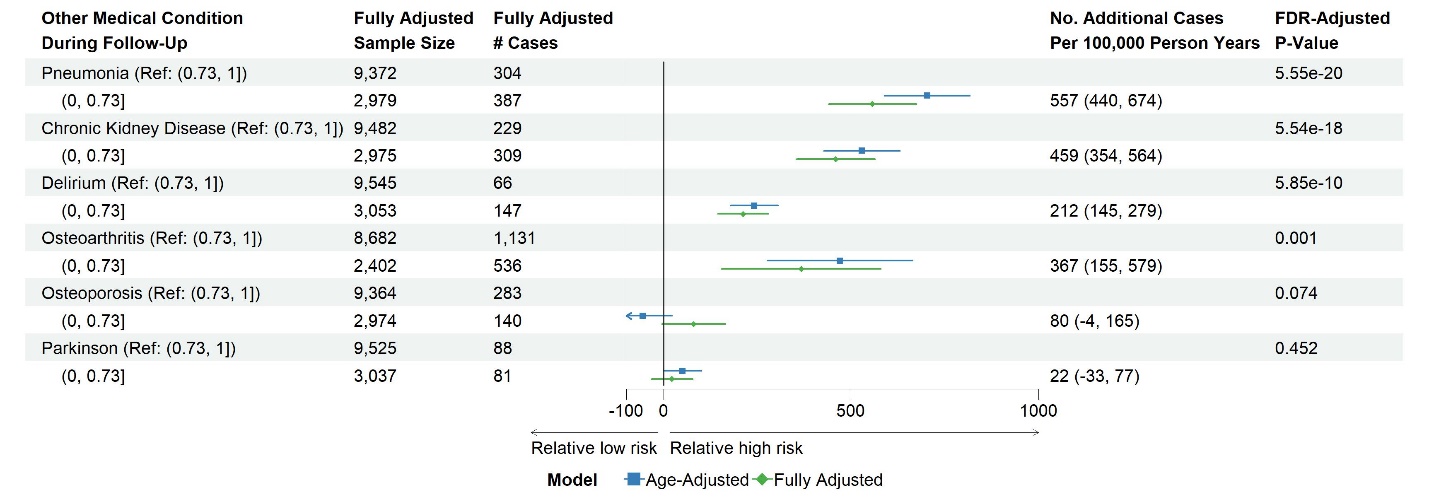

**Figure S8** [UKB] Risk differences comparing low healthspan proteomic score (HPS≤0.73) to high HPS (>0.73) for developing incident medical conditions not in the definition of healthspan (test sample, n=12,935). Fully adjusted covariates: age, sex, ethnicity, BMI, education, Townsend, smoking status, hypertension, hypercholesterolemia, and UKB PPP consortium selection.

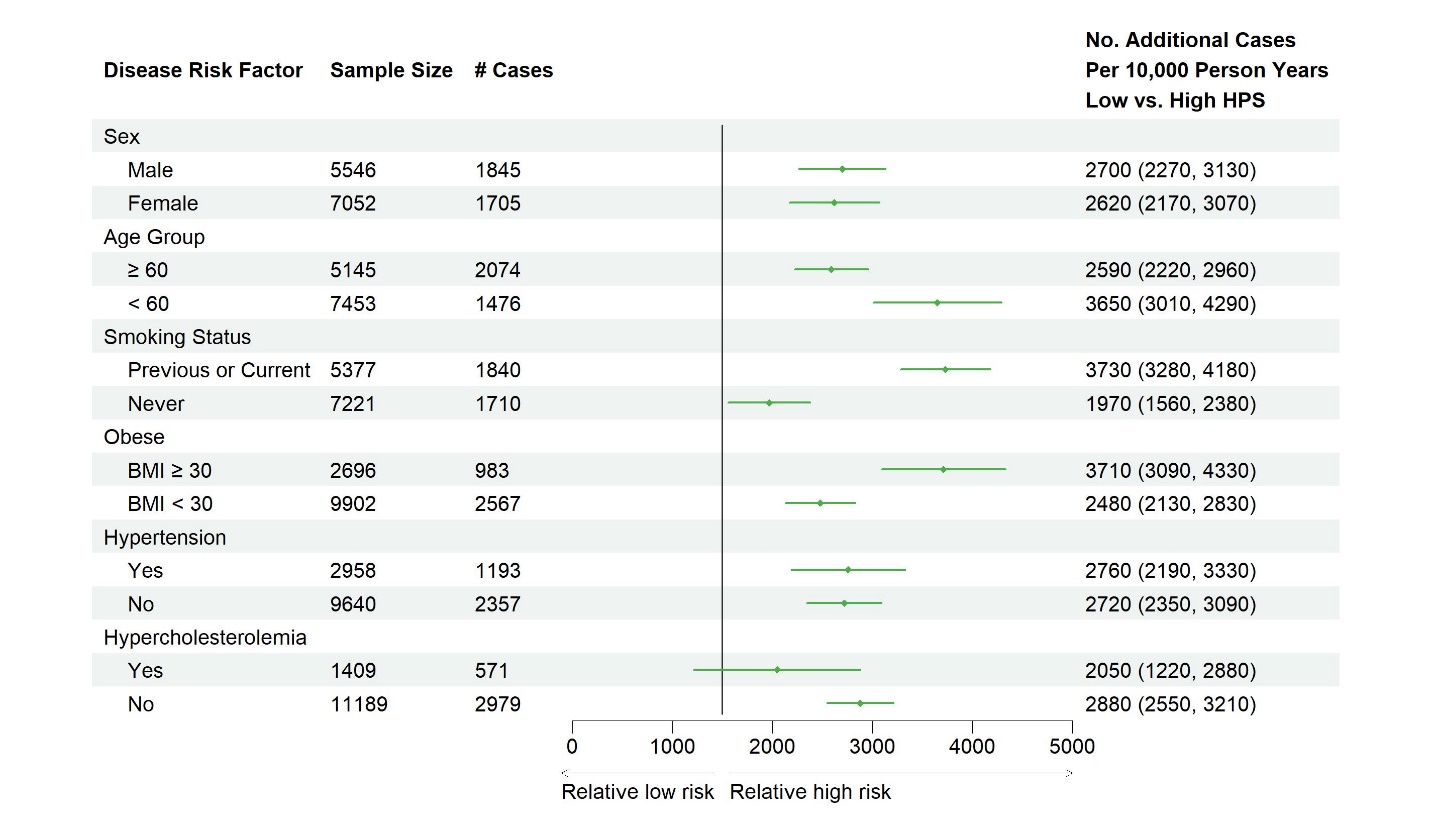

**Figure S9** [UKB] Risk differences comparing low healthspan proteomic score (HPS≤0.73) to high HPS (>0.73) for developing a first condition in the healthspan definition in subgroups (test sample, n=12,935). Fully adjusted covariates: age, sex, ethnicity, BMI, education, Townsend, smoking status, hypertension, hypercholesterolemia, and UKB PPP consortium selection, with the subgroup variable left out. The vertical reference line was placed at the estimate, comparing low HPS to high HPS for developing a first condition in the healthspan definition in all disease-free participants in the test set.

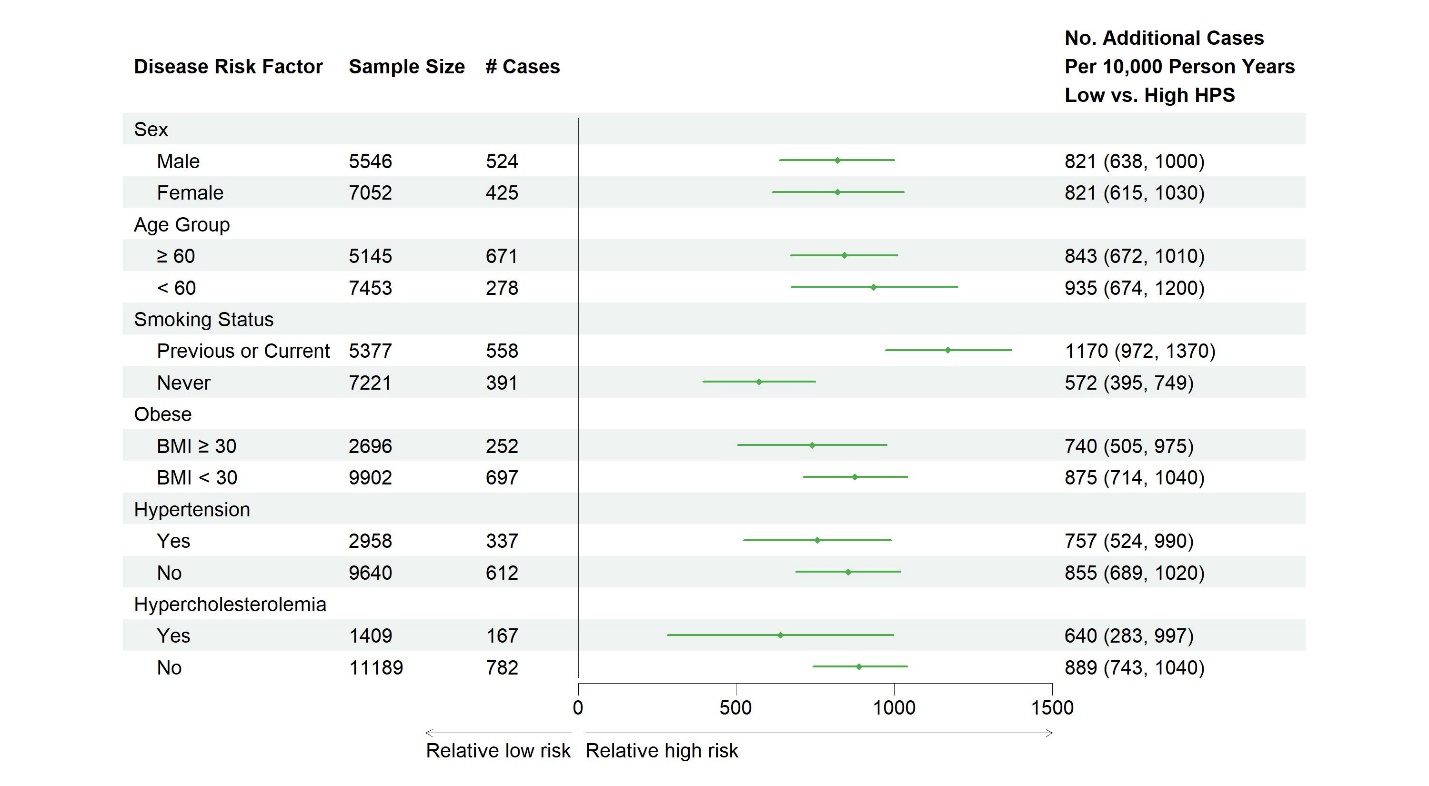

**Figure S10** [UKB] Risk differences comparing low healthspan proteomic score (HPS≤0.73) to high HPS (>0.73) for mortality in subgroups (test sample, n=12,935). Fully adjusted covariates: age, sex, ethnicity, BMI, education, Townsend, smoking status, hypertension, hypercholesterolemia, and UKB PPP consortium selection, with the subgroup variable left out. The vertical reference line was placed at the estimate, comparing low HPS to high HPS for mortality in all disease-free participants in the test set.

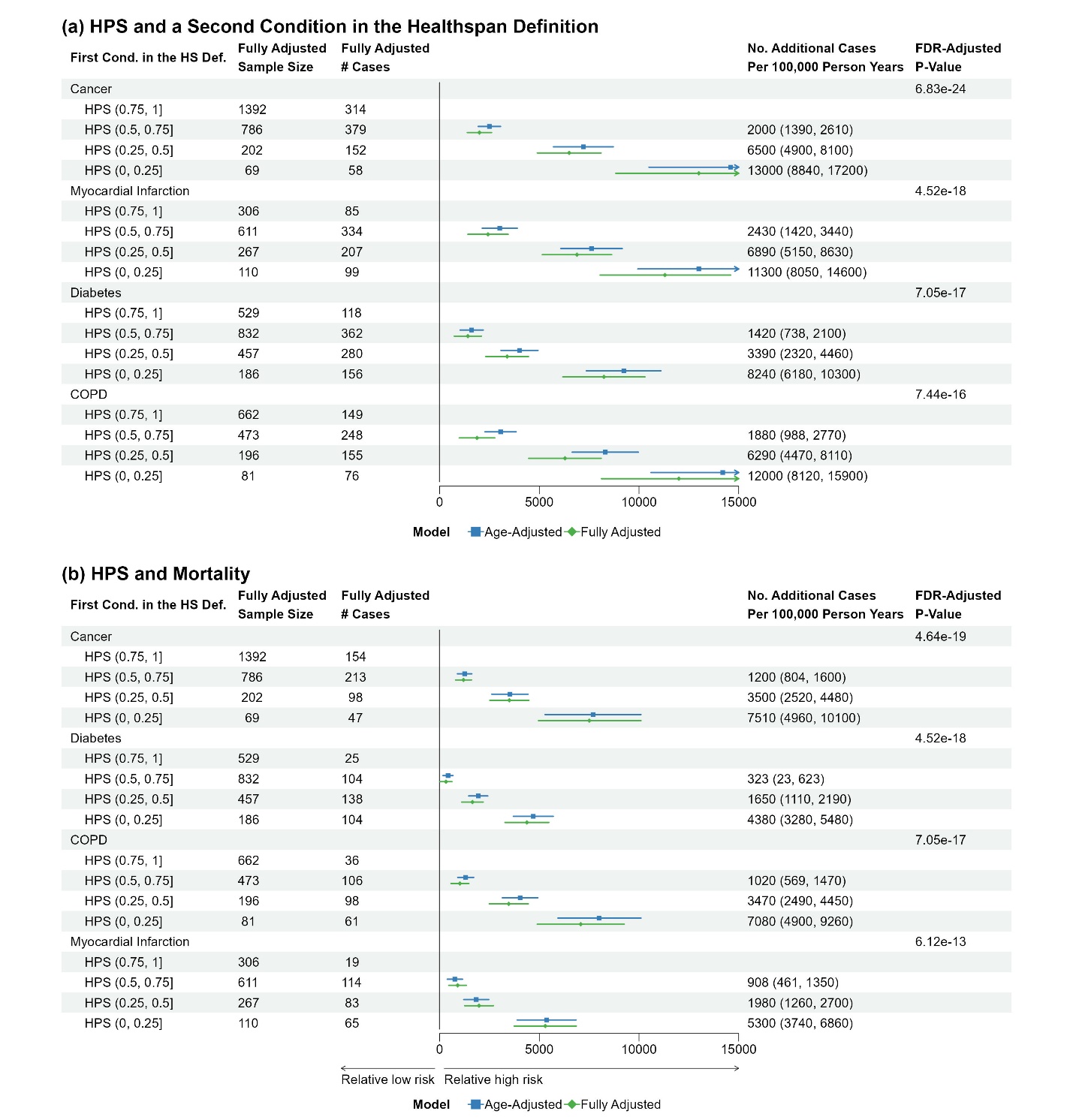

**Figure S11** [UKB] Risk differences comparing healthspan proteomic score (HPS) of (0.5, 0.75], (0.25, 0.5], or (0, 0.25) to HPS (0.75, 1] at baseline for a second condition (a), or mortality (b), in participants initially presenting with a condition in the healthspan definition at baseline (cancer n=2,449, MI n=1,294, diabetes n=2,004, COPD n=1,412). Fully adjusted baseline covariates: age, sex, ethnicity, BMI, education, Townsend, smoking, hypertension, hypercholesterolemia, and UKB PPP consortium selection.

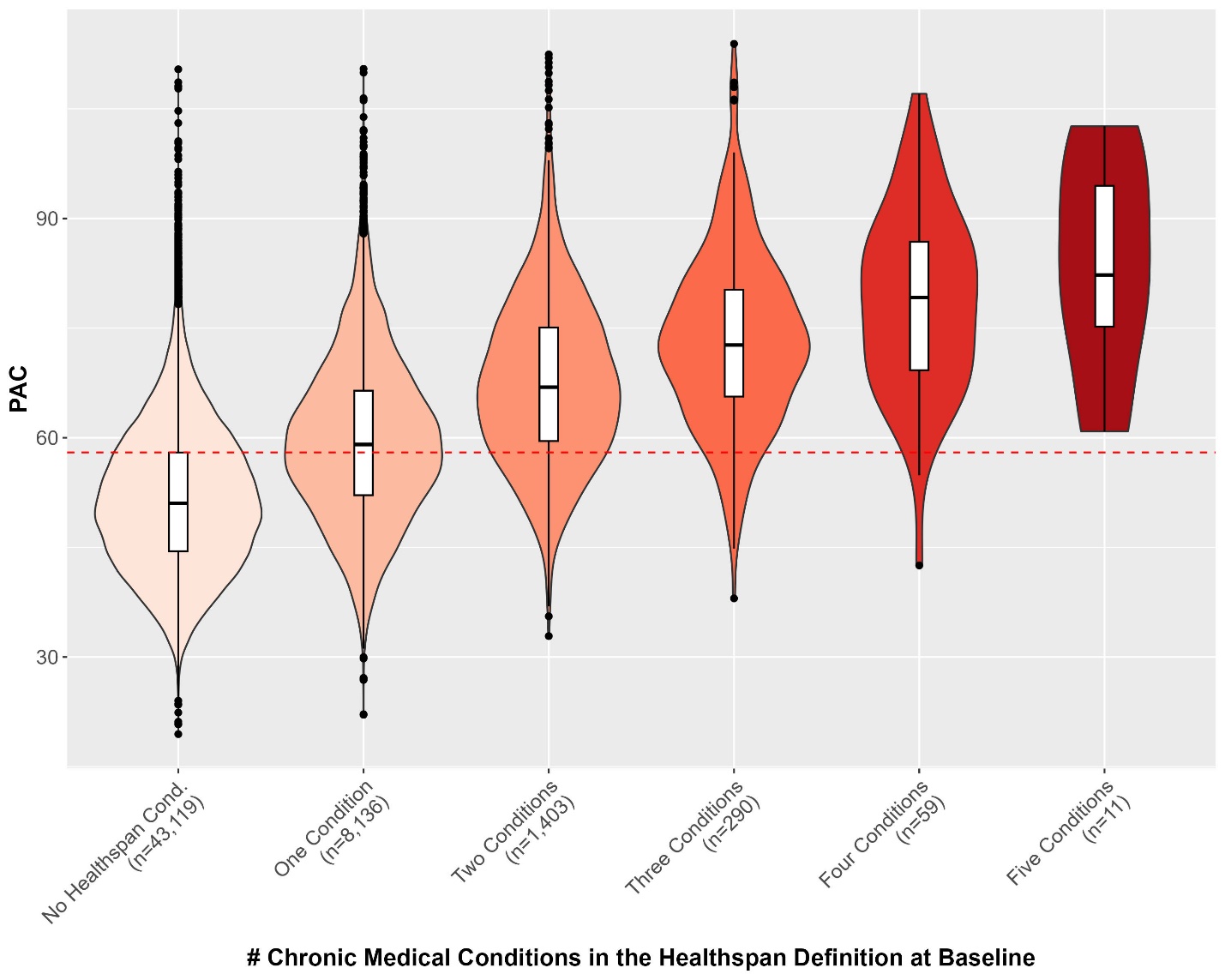

**Figure S12** [UKB] PAC proteomic age distributions in UKB PPP participants (n=53,018), separated by the number of conditions in the healthspan definition at baseline (red dashed line at 58, corresponding to the third quartile)

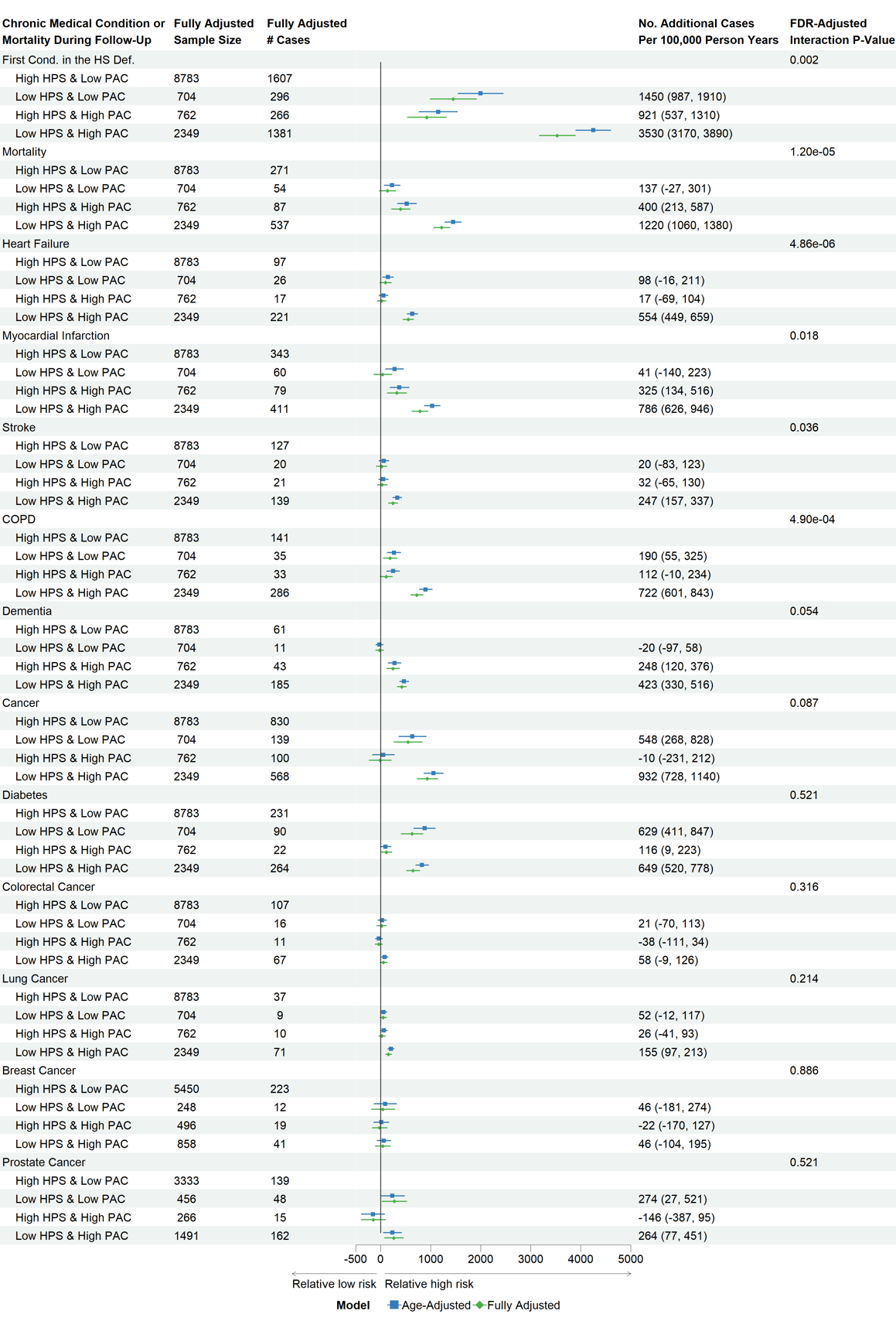

**Figure S13** [UKB] Risk differences comparing more adverse HPS and PAC groups to high HPS and low PAC group for a first condition and individual conditions in the healthspan definition (test sample, n=12,935). Fully adjusted covariates: age, sex, ethnicity, BMI, education, Townsend, smoking, hypertension, hypercholesterolemia, and UKB PPP consortium selection.

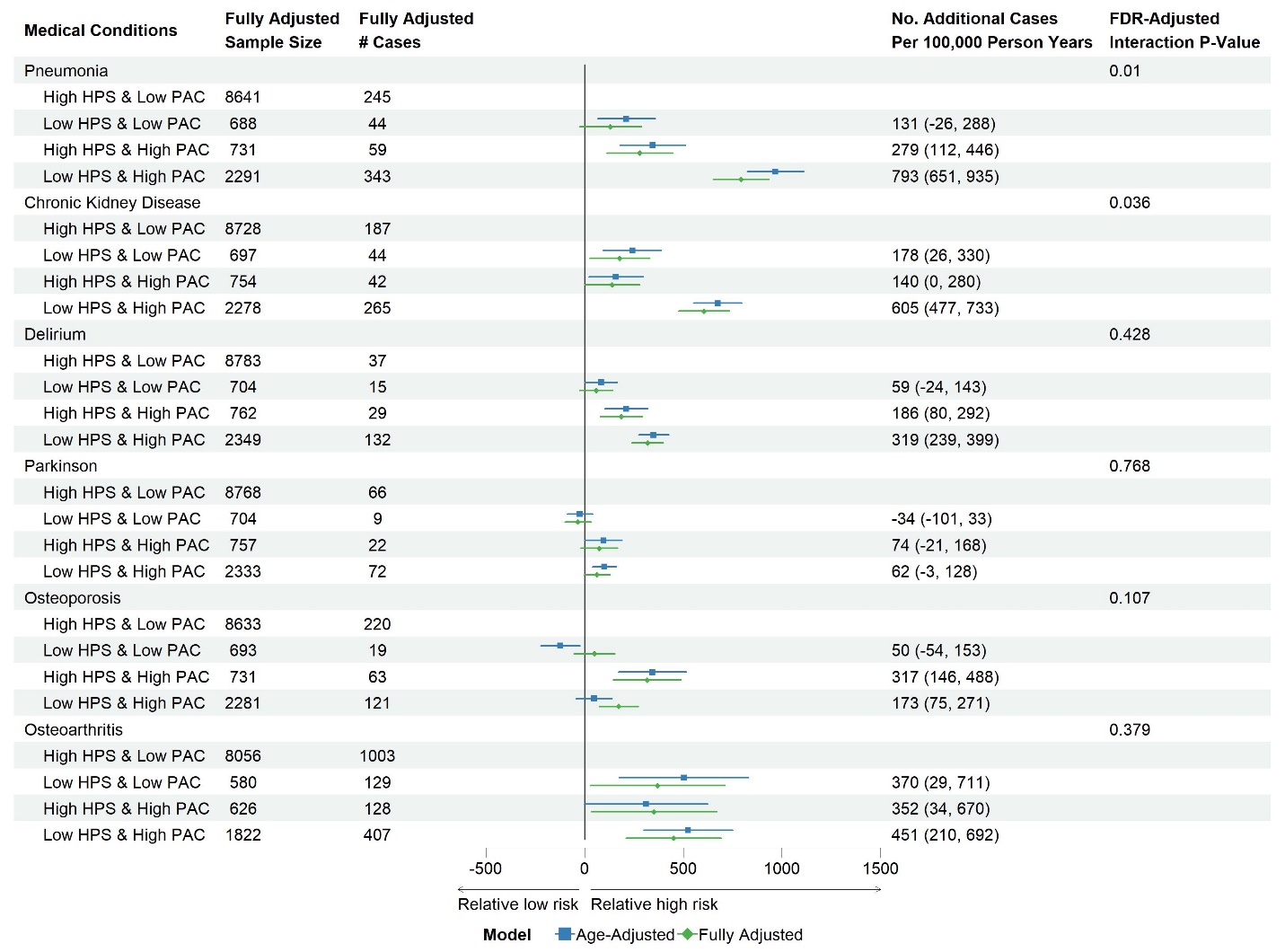

**Figure 14** [UKB] Risk differences comparing more adverse HPS and PAC groups to high HPS and low PAC group for incident medical conditions not in the healthspan definition (test sample, n=12,935). Fully adjusted covariates: age, sex, ethnicity, BMI, education, Townsend, smoking, hypertension, hypercholesterolemia, UKB PPP consortium selection.

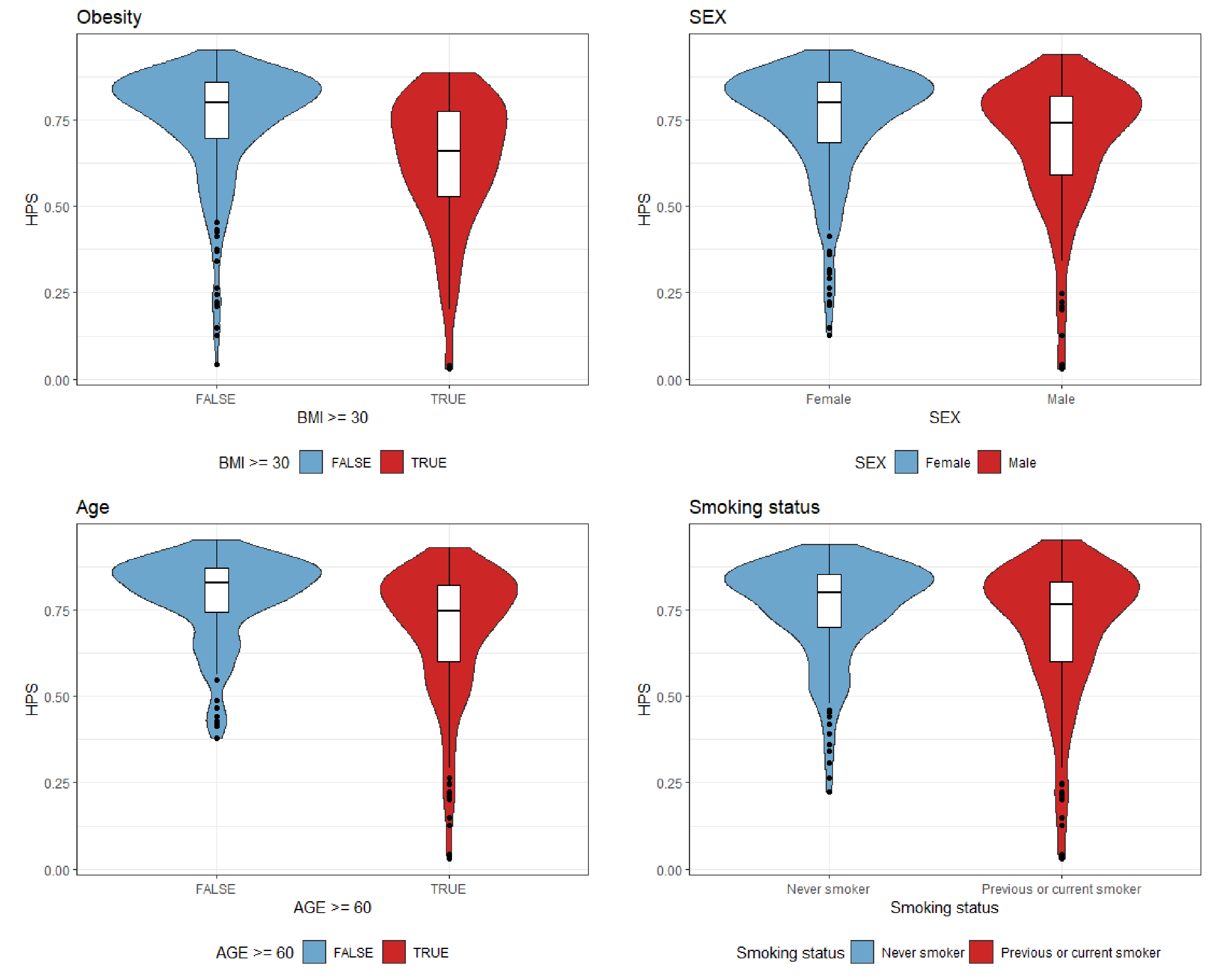

**Figure S15 [EH-Epi] Baseline healthspan proteomic score (HPS) distributions (n=401) comparing (a) males vs. females, (b) older adults vs. younger adults, (c) previous or current smokers vs. non-smokers, (d) obese vs. non-obese participants.**

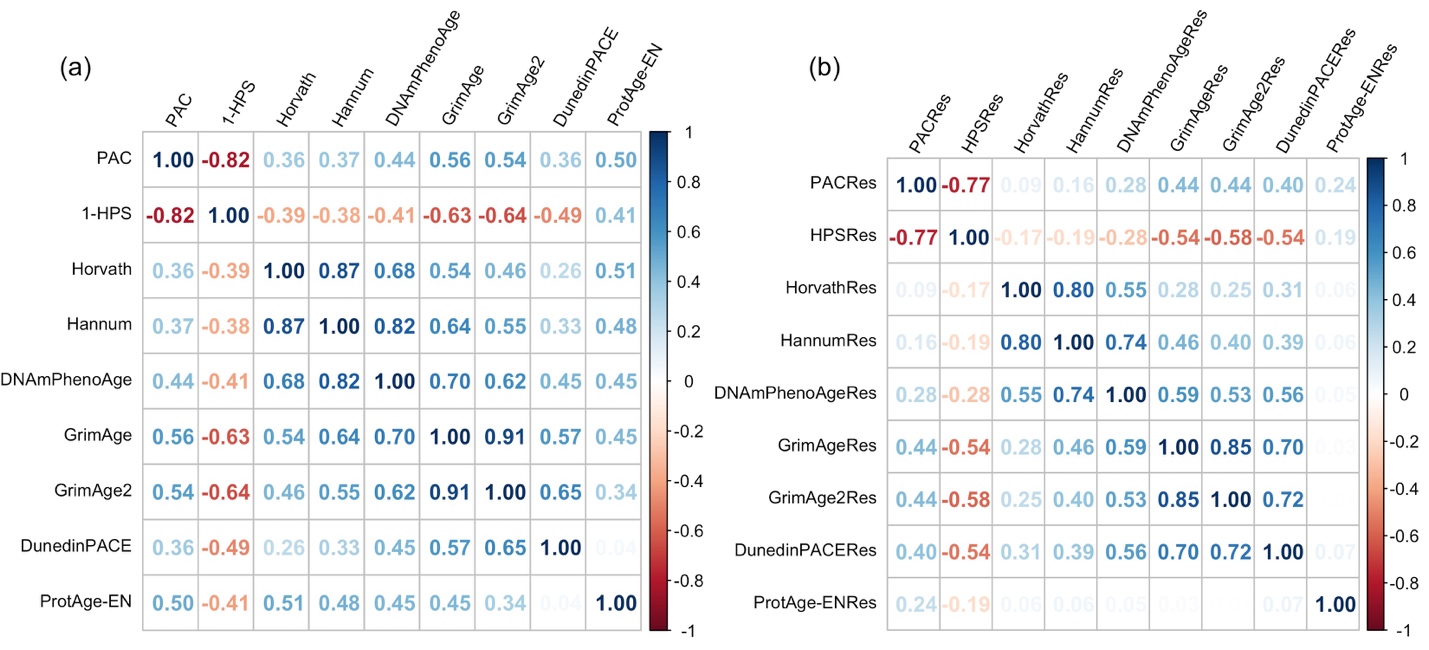

**Figure S16 [EH-Epi] (a) Spearman correlations between HPS (n=401), PAC (n=401), and epigenetic clocks (n=379) and (b) correlations between their residuals after removing the effect of chronological age using linear regression models**

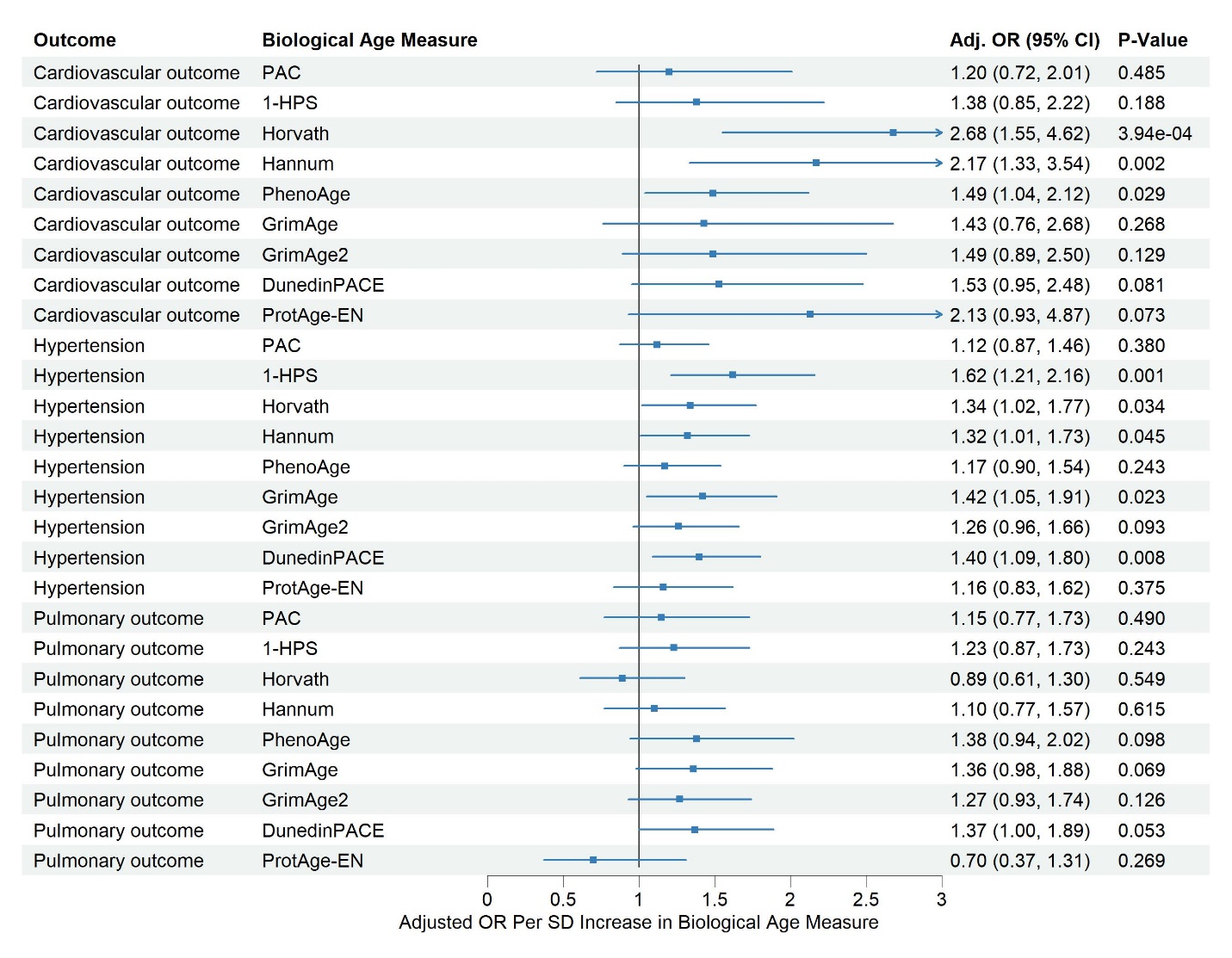

**Figure S17 [EH-Epi] Associations of HPS (n=401), PAC (n=401), and epigenetic clocks (n=399) with hyptertension, cardiovascular, and pulmonary outcomes at the time of blood sampling, adjusted for sex and chronological age.**

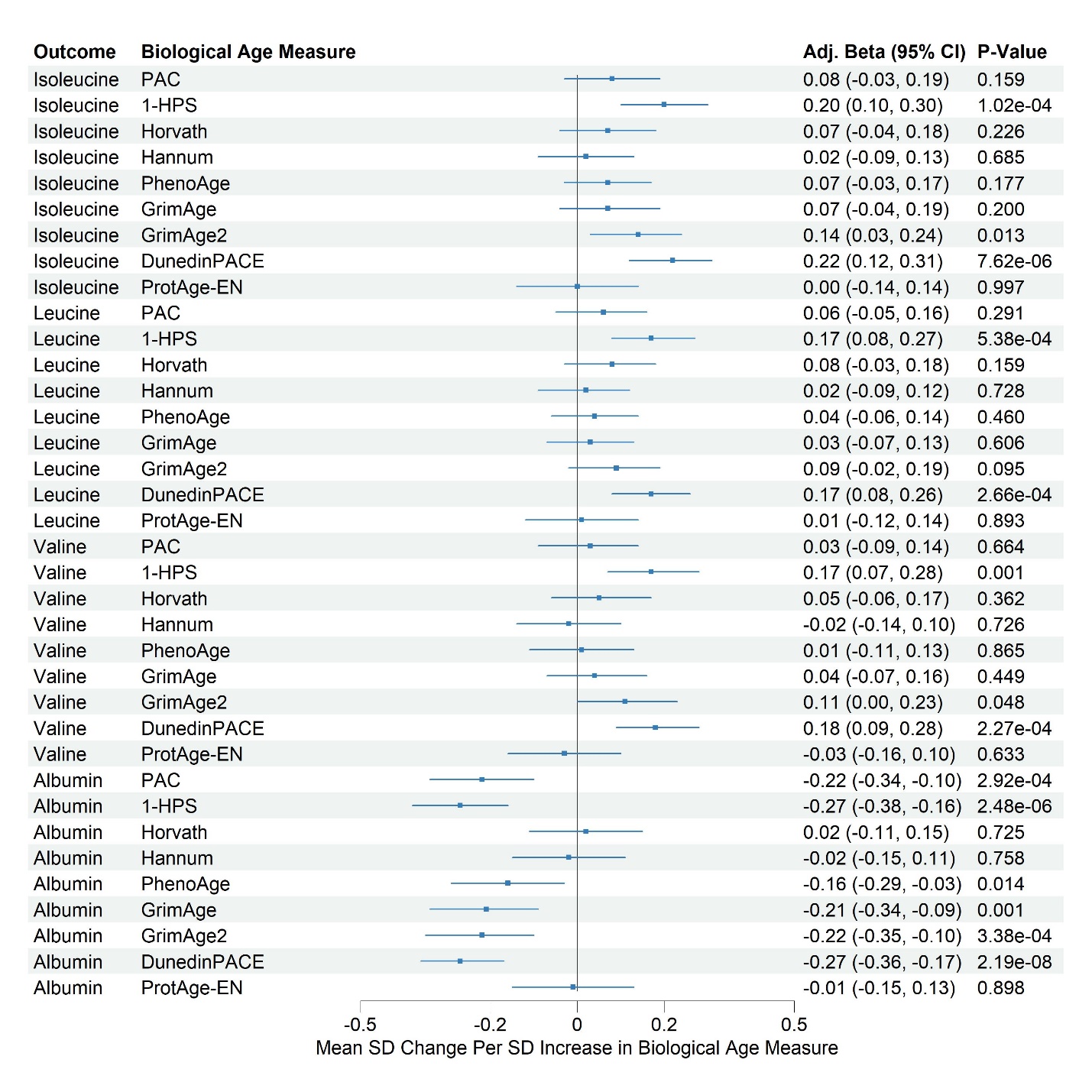

**Figure S18 [EH-Epi] Associations of HPS, PAC, and epigenetic clocks with blood metabolites (n=394) at the time of blood sampling, adjusted for sex and chronological age.**
